# PK-RNN-V E: A Deep Learning Model Approach to Vancomycin Therapeutic Drug Monitoring Using Electronic Health Record Data

**DOI:** 10.1101/2022.05.24.22275271

**Authors:** Masayuki Nigo, Hong Thoai Nga Tran, Ziqian Xie, Han Feng, Bingyu Mao, Laila Rasmy, Hongyu Miao, Degui Zhi

**Affiliations:** Division of Infectious Diseases, Department of Internal Medicine, The University of Texas Health Science Center at Houston, McGovern Medical School, Houston, Texas, United States; Landmark Health, Huntington Beach, California, United States; School of Biomedical Informatics, University of Texas Health Science Center at Houston, Houston, Texas, United States; School of Public Health, University of Texas Health Science Center at Houston, Houston, Texas, United States

**Keywords:** Vancomycin, Recurrent neural network, Pharmacokinetics, Deep Learning, Bayesian model

## Abstract

Vancomycin is a commonly used antimicrobial in hospitals, and therapeutic drug monitoring (TDM) is required to optimize its efficacy and avoid toxicities. Bayesian models are currently recommended to predict the antibiotic levels. These models, however, although using carefully designed lab observations, were often developed in limited patient populations. The increasing availability of electronic health record (EHR) data offers an opportunity to develop TDM models for real-world patient populations. Here, we present a deep learning-based pharmacokinetic prediction model for vancomycin (PK-RNN-V E) using a large EHR dataset of 5,483 patients with 55,336 vancomycin administrations. PK-RNN-V E takes the patient’s real-time sparse and irregular observations and offers dynamic predictions. Our results show that RNN-PK-V E offers a root mean squared error (RMSE) of 5.39 and outperforms the traditional Bayesian model (VTDM model) with an RMSE of 6.29. We believe that PK-RNN-V E can provide a pharmacokinetic model for vancomycin and other antimicrobials that require TDM.

**Statement of Significance:** *Problem:* Current traditional Bayesian models for vancomycin levels were tested in only a limited patient population and take limited patient-specific features. Hence, a more flexible and powerful model, such as deep-learning models, may provide significant advantages.

*What is Already Known:* The Bayesian models do not predict the vancomycin levels well in patient populations with unstable hemodynamic status and fluctuating kidney functions.

*What this Paper Adds:* Deep-learning based pharmacokinetic model for vancomycin (PK-RNN-V E) provided better prediction accuracy with integrating multiple patient-specific features from time sequence electronic health record data. This study proved the concept of model.

## Introduction

Vancomycin is a glycopeptide antibiotic that has been on the market for over 50 years.[1] As the prevalence of Methicillin-resistant *Staphylococcus aureus* (MRSA) has increased since the 1980s, this fundamental antibiotic has become one of the most frequently used in the hospital, especially for the management of severe gram-positive infections.[2] Despite the advent of newer alternative antimicrobials against this pathogen, vancomycin remains a clinically useful agent. Inappropriate vancomycin dosing, however, has been associated with severe side effects, especially nephrotoxicity, therapeutic failure, and the emergence of bacterial resistance. Hence, therapeutic drug monitoring (TDM) of vancomycin levels has been recommended for the use of vancomycin.[3]

Currently, three major approaches to dose individualization are used to estimate pharmacokinetic (PK) parameters: (1) linear regression analysis, (2) population methods, and (3) Bayesian methods.[4] The most recent consensus guidelines from the American Society of Health-System Pharmacists (ASHP), Infectious Diseases Society of America (IDSA), and Society for Infectious Diseases Pharmacists (SIDP) recommend Bayesian-guided dosing for TDM and estimation of area under the curve (AUC) of vancomycin level over a minimal inhibitory concentration (AUC/MIC) of MRSA.[3] Traditional Bayesian models for vancomycin levels (referred to Bayesian Model in this manuscript) use basic patient demographic data with previous population PK data and patient serum creatinine levels to provide the dosing recommendation from the estimation of AUC and the PK parameters. The models provide a more accurate estimation of the AUC/MIC of vancomycin than does the estimation based on the trough concentrations.[4] In addition, Bayesian models estimate the distribution of an individual patient’s PK parameter values based on actual drug serum concentrations, such as vancomycin serum concentrations and subsequent creatinine levels, from the patient (“patient-specific parameters”). After being validated in various patient subpopulations, multiple Bayesian dose-optimizing software programs are available.[5,6]

Despite advances in the models of vancomycin PKs, there are a number of limitations of current Bayesian models. First, each Bayesian model is designed for a specific subgroup of patients based on the population characteristics, which may not cover the diverse real-world patient population.[3,7] Some studies suggest that as many as 58% of real-world patients may be excluded from clinical research.[8] This is further complicated by clinical practice settings, in which there is an interplay among patient-related, disease-related, and system-related factors.[9] Second, these models are often evaluated in patients with stable PK parameters and may not cover critically ill conditions well, in which the volume of distribution and the elimination rates fluctuate acutely.[5,10,11] Finally, these Bayesian methods take only a limited number of patient-specific variables as input, including simple demographics, creatinine levels, vancomycin doses, the infusion time, and vancomycin levels, while there are potentially other relevant patient characteristics, such as other concomitant medications and vital signs, that potentially improve the prediction.[5] Therefore, more powerful and flexible models, such as deep learning models, provide significant advantages, as the models can integrate a wide range of patient-specific features, use flexible time steps, update the model with a local patient population, and cover a wide variety of populations.

Real-world data-driven precision dosing is of great interest to the medical community, as the availability of genetic data and electronic medical records (EHRs) is rapidly expanding.[9,12] In the United States, EHR adaptation has accelerated since the Meaningful Use program was introduced as part of the 2009 Health Information Technology for Economic and Clinical Health Act. EHRs contain both structured and unstructured data, which provide a complete medical history of the patient.[13] Data collection and utilization have been further enhanced with data exchange among different hospital systems or EHR vendors. Artificial intelligence (AI) methods, especially deep learning, have achieved great success in modeling complex EHR data.[14] In particular, sequential models, such as recurrent neural networks (RNNs), enable the modeling of sequential events, which is critical for PK models to provide dynamic changes in PK parameters. Further, RNNs with medical code embedding can take input directly from a real-time EHR data stream, automatically make the adjustments to reflect subtle changes, and provide real-time outputs. Despite the potentially high expressive power of deep learning models, however, no deep learning models for the monitoring of vancomycin levels are available.

The aim of this study is to develop a novel PK approach with RNN-based methods for vancomycin (PK-RNN-V) with EHR data to achieve more accurate and individualized predictions for vancomycin serum concentration in hospitalized patients. Unlike existing RNN models for EHR data for disease risk prediction that have simple output layers, we design a specific PK output layer. To the best of our knowledge, this PK-RNN-V model is the first deep learning-based vancomycin prediction method in the literature.

## Material and Methods

### Data extraction and cohort definition

Our study cohort was extracted from the Memorial Hermann Health System (MHHS) EHR data warehouse. MHHS is one of the largest healthcare systems, consisting of 14 hospitals in the greater Houston area, Texas, United States. The MHHS EHR data warehouse is a rich source of information, as it includes longitudinal information about patients’ demographics, encounters (admission and discharge information), diagnostic codes, laboratory results, vital signs, medication administration, nurse documentation, and other encounter-level administrative data. Patients who were at least 18 years old and who received at least one dose of intravenous vancomycin were identified from the database during the study period of August 2019 to March 2020. For our model training and evaluation, we considered only encounters in which patients had at least one serum vancomycin level after their first vancomycin dose during their hospitalization. We excluded patients who had inappropriate timing of vancomycin levels (e.g., vancomycin level measured during the infusion of vancomycin, the level was measured more than 48 hours after the last dose of vancomycin). We excluded patients who received renal replacement therapy, such as hemodialysis, and extracorporeal membrane oxygenation (ECMO), based on their diagnostic codes (the diagnostic codes are found in the Supplementary Materials) and nurse documentation. To ensure patient data privacy and security during later steps of data handling and modeling, we de-identified the extracted cohort using the methods recommended by U.S. Department of Health and Human Services (DHHS).[15] This study was approved by internal review boards (IRBs) of the University of Texas Health Science Center at Houston and the Memorial Hermann Hospital System.

### Problem setup

Data included in the analysis from EHRs comprised 30 laboratory tests, five vital signs, and 324 types of medications with 911 different medication codes and formulary (Table S7) in addition to encounter and demographic data, such as age at the time of the encounter, gender, ethnicity/race, height, and weight. All of these features were extracted at the encounter level. We used the time of the earliest records of the encounter and the timestamp of the last vancomycin level during the encounter to define the start and stop times for the encounter.

The prediction of vancomycin levels is made at all time points via a PK model based on all of the observed data within the encounter right before the time point. The time step to update parameters of the PK model was defined by the time of vancomycin administration, vancomycin level obtained, or the end of the day; and the data between the time steps were aggregated by the next time step. Because some patients may not receive vancomycin for more than 24 hours, depending on the kidney function or previous vancomycin levels, we used the time step of “the end of the day” to update the model to avoid any information leakage. Figure 1A displays a simplified encounter in which patients have multiple vancomycin doses and levels and other lab tests (e.g., creatinine levels) taken during the windows previously defined. Using this time-step scheme, we can make the relationship between the dose and the levels more explicit in terms of the model while not losing information during days in which patients did not have vancomycin administrations or levels. We also can make the relationship more practical and provide real-time predictions in regard to the drug concentrations. In MHHS, the infusion time of vancomycin varies, depending on the dose of vancomycin; ≤1000 mg: infuse over 1 hour, 1001–1500mg: infuse over 1.5 hours, 1501–2000mg: infuse over 2 hours, and >2000mg: infuse over 2.5 hours. To simplify the model, we used 1 gm/hour for the infusion time. We further assume that vancomycin measurement should not be taken during the infusion. Missing values are imputed with previous values, assuming that clinicians, due to patient stability, did not repeat the tests. When the values have never been measured for the patient, the values were imputed using the mean value. The continuous values are standardized to *z*-scores.

**Figure 1:**
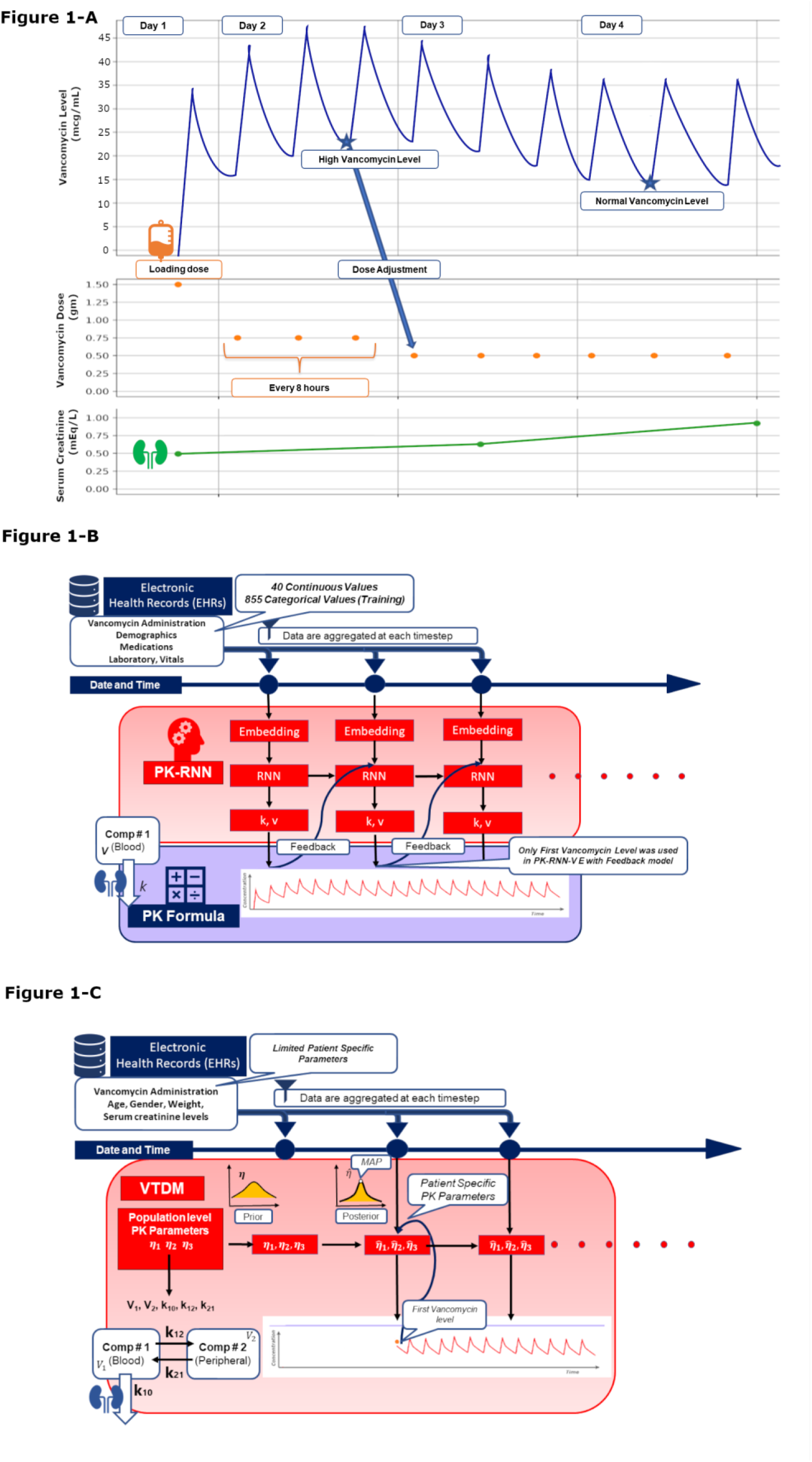
Simplified Example Patient and PK-RNN-V Model Architecture and k: Elimination of Vancomycin Rate from The Compartment, Comp: Compartment, MAP: Maximum A Posteriori, PK: Pharmacokinetic, RNN: Recurrent Neural Network, v: volume distribution of vancomycin, VTDM: Vancomycin Therapeutic Drug Monitoring (Traditional Bayesian Model)

**Figure 1A**. Simple schematic time sequence of an example patient from our cohort. Vancomycin serum concentration is demonstrated in the upper figure. The blue dash line represents the predicted vancomycin serum concentration in the patient. The blue stars indicate the actual vancomycin levels. In the middle graph, orange dots are vancomycin doses, and the green line in the lower figure is the creatinine level. The vancomycin levels or other key measurements can be irregular, depending on the dosing intervals or patient clinical status. **Figure 1B**. Schematic structure of the PK-RNN-V model. PK-RNN-V model is one compartment model. Vancomycin doses, basic demographics, laboratory, concomitant medications, and vitals were extracted from EHRs. All data were aggregated at the time step, and the data were fed into the models. RNN updates *k* (vancomycin elimination rate) and *v* (volume distribution of vancomycin). These two parameters and infusion data (dose and timing) were used to generate vancomycin pharmacokinetic (PK) curves with the PK formula. Feedback models use the first observed vancomycin level at the inference time to update the model parameters for predicting the vancomycin level after the first observation. **Figure 1C**. Schematic structure of VTDM model. VTDM model is a two compartment model, and only takes the aggregated data of vancomycin administration, age, gender, weight, height, and serum creatinine from EHR to update PK parameters. The initial parameters (*η*_1,_ *η*_2_, *η*_3_) are provided from previous studies as population based PK parameters. VTDM with feedback model uses the first vancomycin level to adjust those parameters as maximum a posteriori (patient-specific parameters). This provides *v*_*1*_, *k*_*10*_, *k*_*12*_, and *k*_*21*_ of the patient to predict vancomycin concentration.

### Model Design

#### PK-RNN-V and PK-RNN-V E models

We reason that drug-level predictions in real-world data need to meet the following four requirements: (i) use demographic and irregular historical EHR data as input, (ii) take observations of the drug levels or relevant physiological biomarkers at irregular time intervals, (iii) use the drug levels that follow underlying PK models with varying parameters, and (iv) re-estimate PK parameters each time an observation is taken. Based on these requirements, we design an autoregressive RNN model with a PK prediction head whose predicted vancomycin can be evaluated at arbitrary time points (Figure 1B). Autoregressive architecture is commonly used in neural machine translation models and generative models, such as GPT-3 [16,17], to capture context and temporal correlation. In addition, we use RNN, as it is a natural choice for modeling longitudinal EHR data.[14,18–20] We have shown that RNN can successfully model heart failure risks.[21] For drug levels, we predict the underlying PK parameters directly from the RNN; then, vancomycin levels at any time point can be calculated automatically by the PK equation. (Figure 1A) For simplicity, we use a one-compartment model. Irregular time intervals between observations can be modeled by PK calculations.

The overall architecture of the PK-RNN-V model has three components: (i) a code embedding layer to take inputs from each time step, (ii) an RNN layer to update the hidden representation and output the elimination rate of vancomycin (k) and the volume of distribution (v), and (iii) a one-compartment PK model to calculate the vancomycin concentration. The information for each patient is represented by a sequence of continuous and categorical variables. There are 40 continuous variables in our model, including selected laboratory tests, vancomycin dosing, and demographic information (a detailed description of the PK-RNN-V model is found in the Supplementary Materials).

The categorical variable contains all medication codes. The medication codes are embedded into 8-dimensional vectors and then summed across each event. The embedding layer was initialized randomly, where the weights were drawn from a standard Gaussian distribution. No diagnosis codes are used in this model because we found, empirically, that adding diagnosis codes to the model does not improve the performance. The input to the RNN layer is a 48-dimensional vector (40+8) at each time step, consisting of the medication embedding (8) and the normalized continuous variables (40). The RNN layer is a single layer GRU with a hidden size of 32. The output layer is a linear layer of size (32, 2), which maps the hidden layer of GRU to the volume of distribution V and the elimination k. Since k and V should be nonnegative, the exp nonlinearity was added on top of the linear layer to get the desired output. For the PK model, the total amount of vancomycin in the patient system is calculated based on the following differential equation:

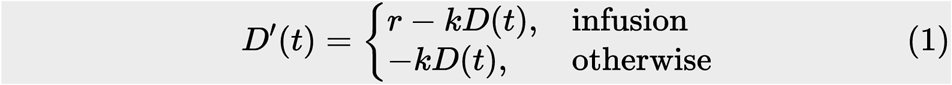

*D* is the mass of vancomycin in the body, *r* is the infusion rate, *k* is the elimination rate).[22,23] Solving this equation, we have:

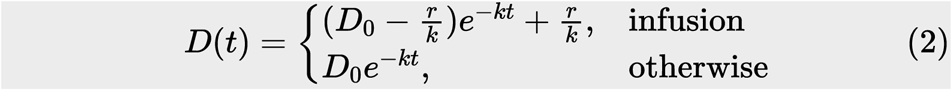

We use a mean squared error as the loss function to train our model. In addition, we add two regularizations. We have a baseline elimination rate *k* and a baseline volume of distribution *v*. The baseline elimination rate *k* is calculated from the Matzke equation *k* = 8.3e-4 * CrCl (creatinine clearance in mL/min) + 4.4e-3, in which the creatinine clearance is calculated from the Cockcroft-Gault equation (the formulas are found in the Supplementary Materials).[24] The baseline volume of distribution *v* is calculated from the weight as 0.7L/kg * weight (kg). One regularization term penalizes the deviation from the predicted *k* and *v* values from the baseline values. The other regularization term penalizes the sudden change in the predicted *k* and *v* values, using the L2 norm of the first-order difference. To train the model, we use adadelta as the optimizer, with a learning rate of 5e-2, epsilon of 1e-4, and weight decay of 0.2. The training minibatch size is set at 50, and we use early stopping with patience of 2 to avoid overfitting.

Several variants of the PK-RNN-V were considered. First, we make the model autoregressive by feeding the predicted vancomycin concentration level back to the input when there is no measurement available and take the true measurement as input otherwise. The intention is to make the model learn to adjust the prediction based on the measurement. To have a fair comparison with VTDM, one variant of PK-RNN-V uses only the first measurement to adjust its hidden state; this variant is termed PK-RNN-V feedback. We also named the variant that takes all measurements available as PK-RNN-V full feedback. We then did an ablation study by excluding the kidney function-related features, such as serum creatinine concentration and glomerular filtration rates, during the training. The trained model is called PK-RNN-V without Kidney Function.

In addition, we use model ensembling to obtain a posterior estimate of the vancomycin concentration. Ten models were randomly initialized and trained using the same dataset and procedure as described above. The prior distribution of the models is uniform. The posterior distribution is adjusted according to the first measurement in each encounter: *P*(*model*|*measurement*) ∝ *P*(*model*)*P*(*measurement*|*model*), where *P*(model) equals 1/10 and *P*(measurement|model) is set to be a Gaussian around the predicted value with unit variance. The final prediction result is then the weighted average of all 10 models, using the posterior. The ensemble model is named PK-RNN-V E. All of the ensembles of PK-RNN-V variants are named similarly by having the suffix E after the PK-RNN-V. The source code of our models is publicly available to enable its application and further evaluation by other researchers. (https://github.com/ZhiGroup/PK-RNN)

### Bayesian model (Vancomycin therapeutic drug monitoring, VTDM)

To evaluate the performance of our proposed model, a Bayesian vancomycin therapeutic drug monitoring (VTDM) model is applied as the baseline.[25] The VTDM model is a population-level Bayesian model built on a two-compartment PK model while adjusting for basic patient information, such as age, gender, weight, and creatinine levels (a detailed description of the VTDM model is found in the Supplementary Materials). The equations and parameter settings are provided in Lim,[25] and the corresponding R code is available on GitHub at https://github.com/asancpt/shiny-vtdm.[26] In addition to the baseline VTDM model, we tested a VTDM feedback model that can adjust the PK parameters according to the first measurements of the vancomycin level. This is based on the consideration that the original VTDM model primarily set the values of volume distribution (*v*) and vancomycin elimination (*k)* based on the population mean of a group of Korean patients,[25] When directly utilizing such a model to predict temporal changes among certain patients, the overlooked individual-level information, the *v* and *k* of such patients, for example, may lead to large prediction bias. VTDM feedback uses a gradient descent to adjust the parameters and to find the maximum a posteriori estimation. By doing so, the original VTDM model is transferred from the population level to the individual level, which is more consistent with the current Bayesian models available in the community.

### Model Evaluation

Data were split into training, validation, and test sets with a ratio of 70:15:15 based on patient identification. We avoided using an encounter ID to assign the patients, as it would potentially assign the same patients to different datasets with different encounter identifications. We used root mean square error (RMSE), mean absolute error (MAE), and mean absolute percentage error (MAPE) to compare the model performance between PK-RNN-V and VTDM models (the formulas are found in the Supplementary Materials). Because the PK-RNN-V E with feedback and VTDM with feedback models used the first vancomycin level to adjust the model parameters, the first measurement was excluded when comparing these models. PK-RNN-V E with the full feedback model used subsequent vancomycin levels to adjust the model. For subgroups analysis, demographic data, ICD-10, ICD-9, and norepinephrine in the medication table were used to identify the subgroups with underlying comorbidities. Our dataset did not have the patient hospital bed information where the patient existed, including whether the patient was in an intensive care unit (ICU). Administration of norepinephrine was used to surrogate ICU status of patients as this medication often only used in ICU. The PK-RNN-V E with feedback model was statistically compared to the VTDM with feedback model by a paired *t*-test. The area under the curve of vancomycin levels over minimal inhibitory concentration (AUC/MIC) was calculated only for PK-RNN-V models due to the absence of a gold standard measurement in our dataset. Finally, the estimated vancomycin levels in the selected patients were depicted based on the predicted *v* and *k*. Python version 3.6 (Python Software Foundation) and PyTorch-1.0.0 were used.

## Results

A total of 12,258 patients with 195,140 encounters were identified from the database during the study period. After the exclusion of 6,775 patients, 5,483 patients with 9,504 encounters in which the patient received at least a dose of vancomycin followed by vancomycin levels were included in our study. Table 1 provides the characteristics of patients included in the study. The median age of the population was 61 years, with a slight majority of males (55%). Race/ethnicity included White (36.4%), African American (19.5%), Hispanic (15%), and Asian (1.5%). Median weight and height were 82.9 kg and 172 cm, respectively. Hypertension (20.1%) and diabetes (11.7%) were common diagnoses among the cohort. A total of 55,336 doses of vancomycin (5.8 doses per encounter) were administered, with a median dosage of 1.0 gm (IQR: 1.0–1.5 gm). Vancomycin levels were measured 18,588 times (1.9 measurements per encounter) at various times after vancomycin initiation. The median level was 14.7 mcg/mL (Figure S1, histogram of vancomycin levels).

**Table 1.**
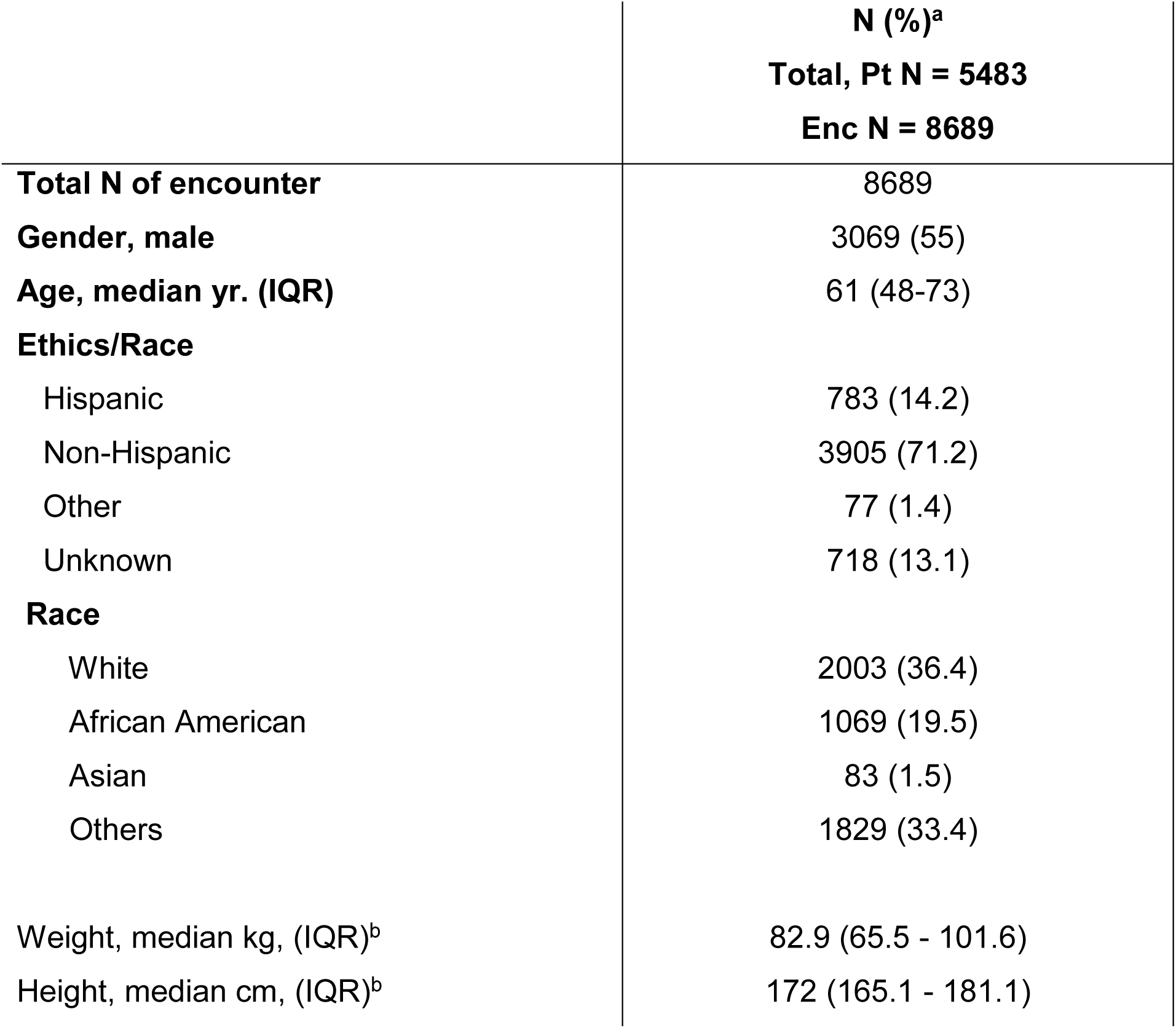
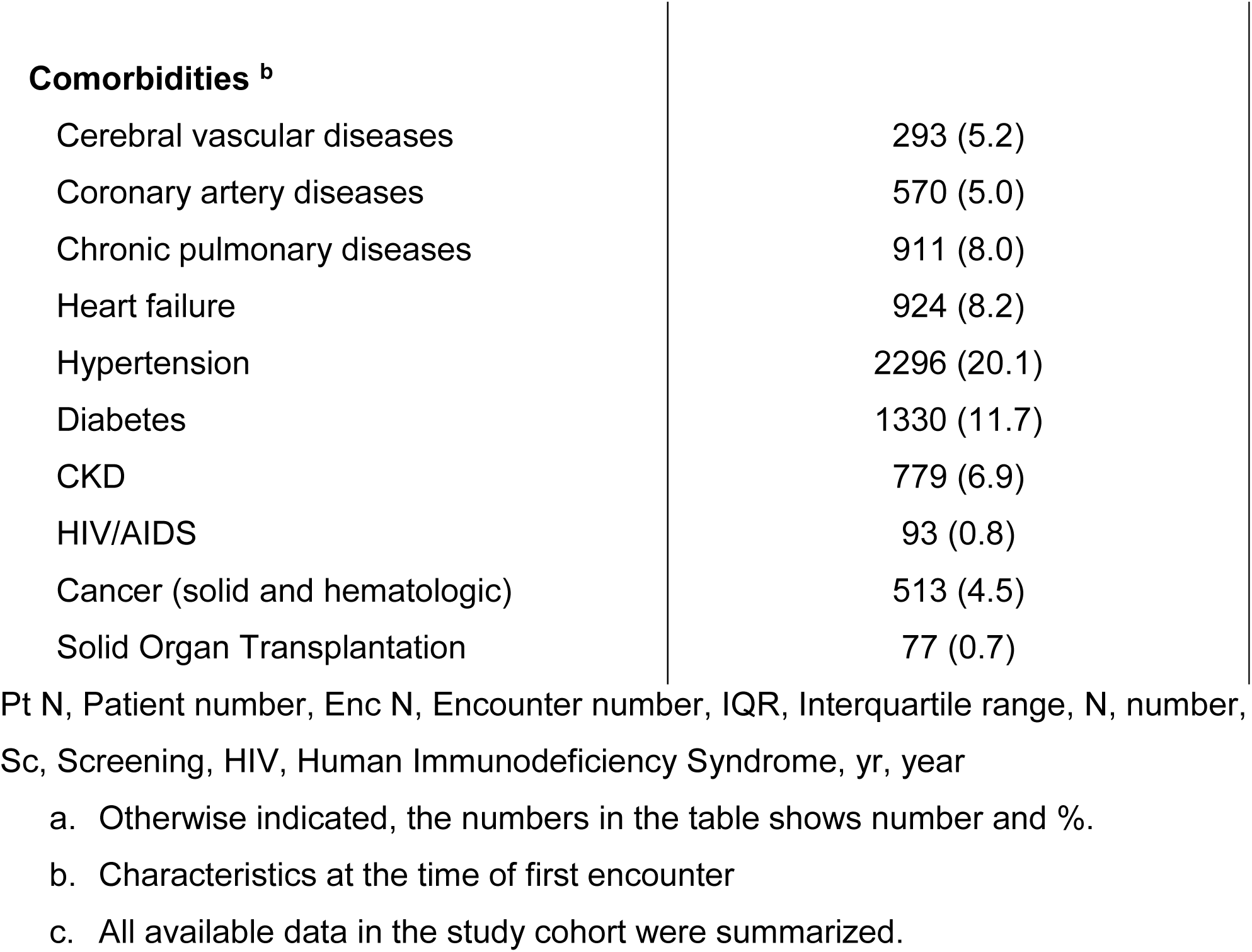
Baseline Characteristics of Patients Included in This Study^*^.

|  | **N (%)<sup>a</sup>**<br>**Total, Pt N = 5483**<br>**Enc N = 8689** |
| --- | --- |
| **Total N of encounter** | 8689 |
| **Gender, male** | 3069 (55) |
| **Age, median yr. (IQR)** | 61 (48-73) |
| **Ethnicity/Race** |  |
| Hispanic | 783 (14.2) |
| Non-Hispanic | 3905 (71.2) |
| Other | 77 (1.4) |
| Unknown | 718 (13.1) |
| **Race** |  |
| White | 2003 (36.4) |
| African American | 1069 (19.5) |
| Asian | 83 (1.5) |
| Others | 1829 (33.4) |
| Weight, median kg, (IQR)<sup>b</sup> | 82.9 (65.5 - 101.6) |
| Height, median cm, (IQR)<sup>b</sup> | 172 (165.1 - 181.1) |
| **Comorbidities<sup>b</sup>** |  |
| Cerebral vascular diseases | 293 (5.2) |
| Coronary artery diseases | 570 (5.0) |
| Chronic pulmonary diseases | 911 (8.0) |
| Heart failure | 924 (8.2) |
| Hypertension | 2296 (20.1) |
| Diabetes | 1330 (11.7) |
| CKD | 779 (6.9) |
| HIV/AIDS | 93 (0.8) |
| Cancer (solid and hematologic) | 513 (4.5) |
| Solid Organ Transplantation | 77 (0.7) |

Pt N, Patient number, Enc N, Encounter number, IQR, Interquartile range, N, number, Sc, Screening, HIV, Human Immunodeficiency Syndrome, yr, year
- Otherwise indicated, the numbers in the table shows number and %. - Characteristics at the time of first encounter - All available data in the study cohort were summarized.

Vital signs and laboratory results throughout the encounters are presented in Table 2. Median temperature was 36.8°C, and median systolic, diastolic, and mean blood pressure were 125 mmHg (interquartile range [IQR]: 112–140), 66 mmHg (IQR: 57–75), and 88 mmHg (IQR: 79–97), respectively. Of note, systolic and diastolic blood pressure measurements had a significant proportion of missing values (62.6% and 87.5%, respectively). Median creatinine level was 0.99 mEq/L (IQR: 0.71–1.44). Total protein and albumin levels were 6.8 g/dL and 2.6 g/dL, respectively. Median white blood cell count was 10.2 k/cm^2^, with an absolute neutrophil count of 7.1 k/cm^2^.

**Table 2.**
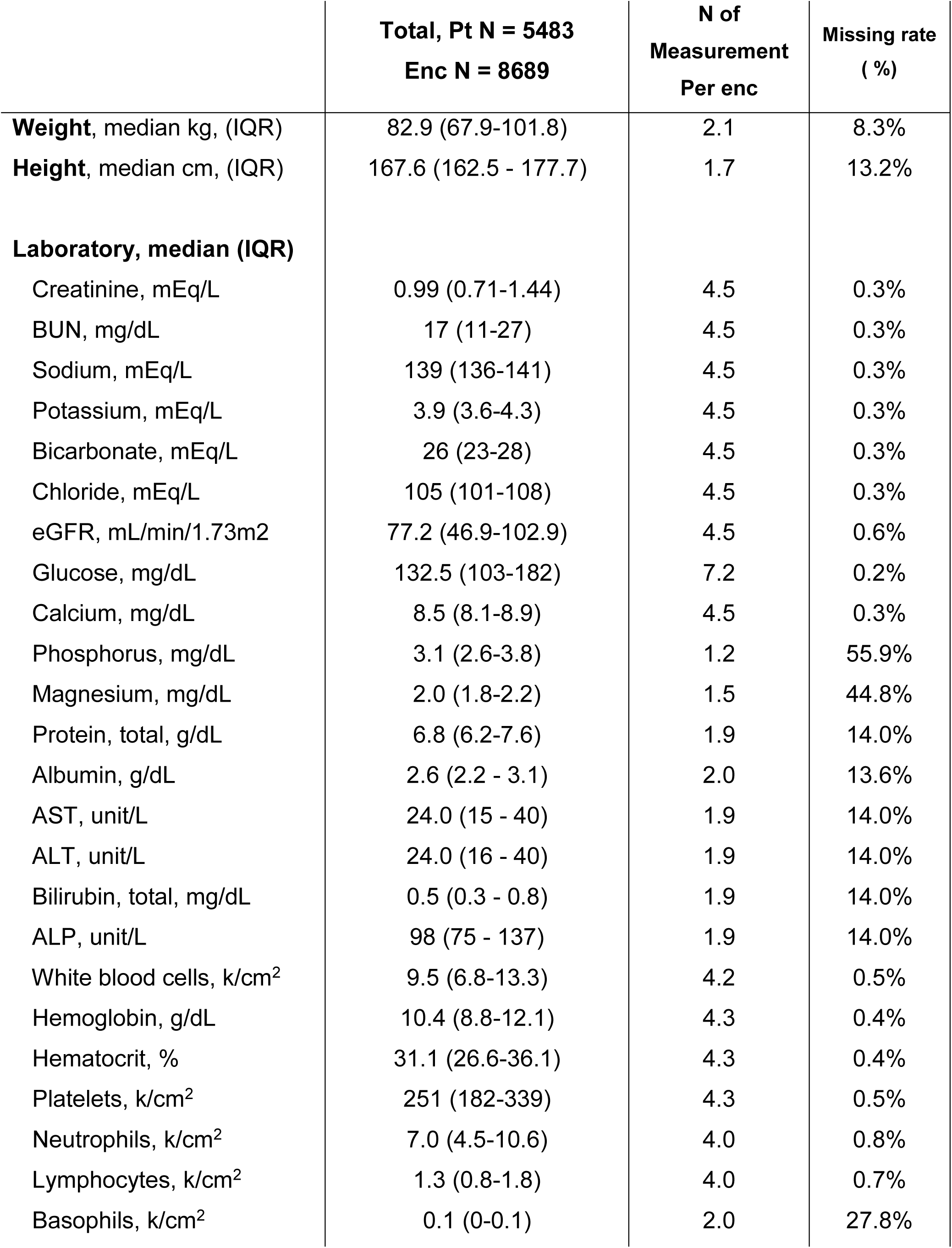
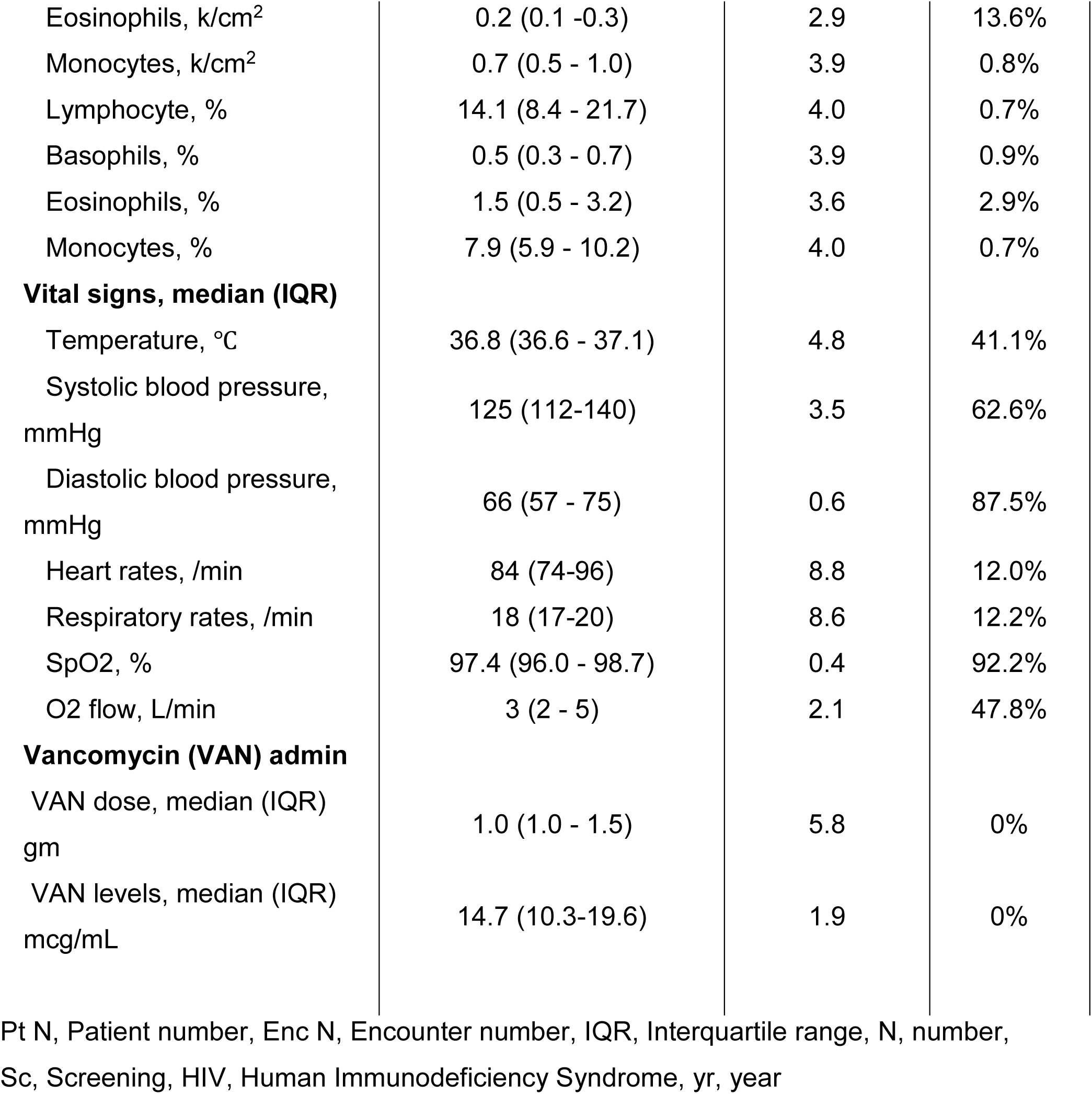
Characteristics of Repeated Measurement Included into the Models.

Table 3 presents the performance of variations of the PK-RNN-V and VTDM models. Notably, all PK-RNN-V models exhibited better RMSE, MAE, and MAPE compared to any of the VTDM models. The baseline PK-RNN-V shows good performance. For the PK-RNN-V model, RMSE = 5.86, MAE = 4.09, and MAPE = 37.57; for the VTDM model: RMSE = 8.58, MAE = 6.54, and MAPE = 41.81. In addition, we showed that the performance of the model was improved by ensembling and letting the model adjust its state based on the first measurement. Statistical comparison (paired *t*-tests) between PK-RNN-V E with feedback and VTDM with feedback revealed RMSE = 5.39 vs. 62.9 (*p*-value = 2.51x 10^−8^), MAE =3.64 vs. 4.26 (*p*-value = 2.51 × 10^−8^), and MAPE = 25.41 vs. 29.15 (*p*-value = 0.00026), respectively. Finally, PK-RNN-V E with full feedback used all of the available measurements to adjust its state and achieved the best result: RMSE = 5.37, MAE = 3.62, and MAPE = 25.05. Interestingly, PK-RNN-V E models performed well even when some of the critical features, such as serum creatinine levels and glomerular filtration rates (GFRs), were masked to the models. We did this ablation study to show that the trained model is able to take other available lab tests or medications to infer the kidney function. The RMSE of the PK-RNN-V E model without creatinine/GFR is 5.91 compared to 6.29 in the VTDM feedback model (Table S1).

**Table 3.**
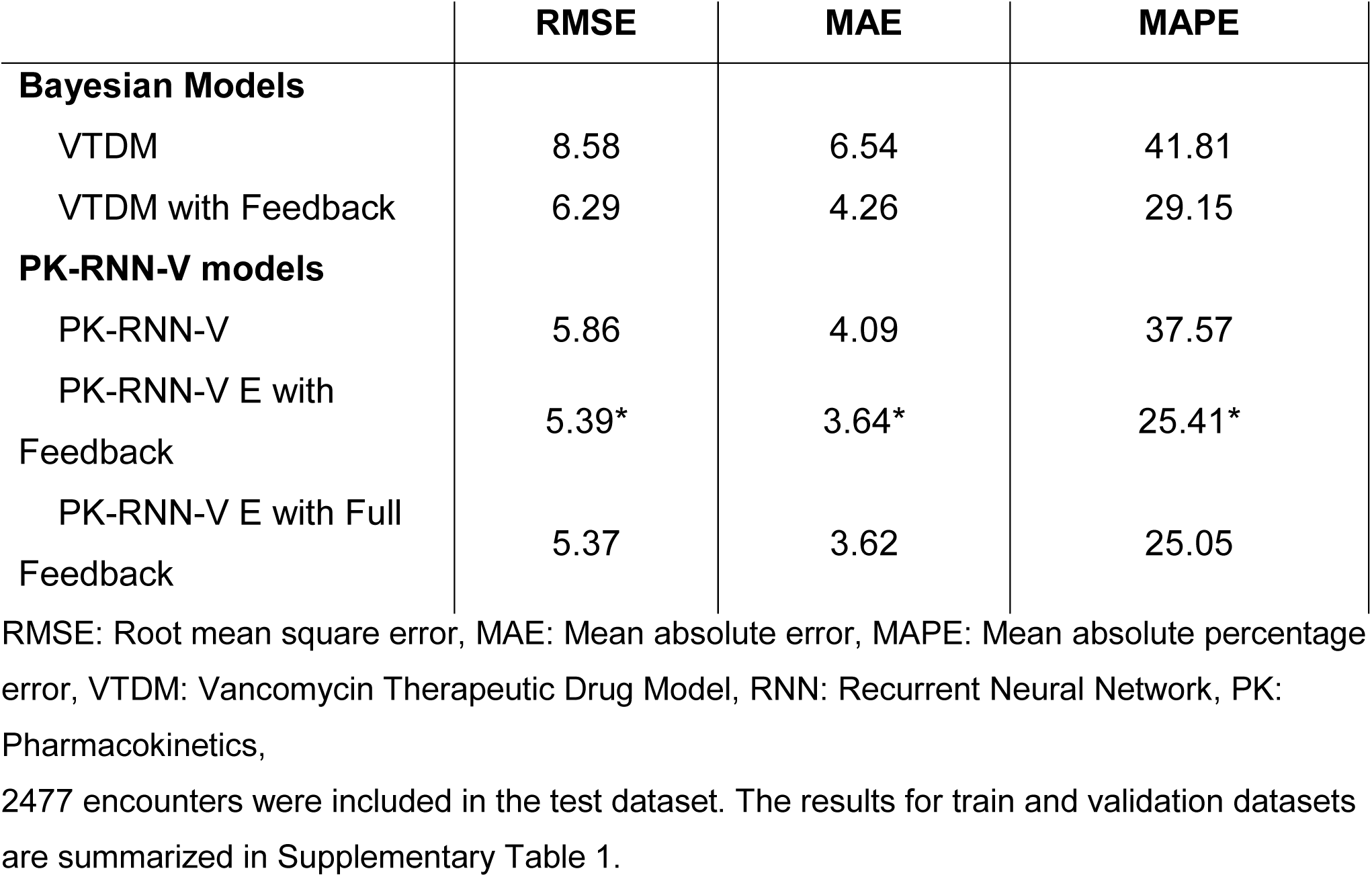
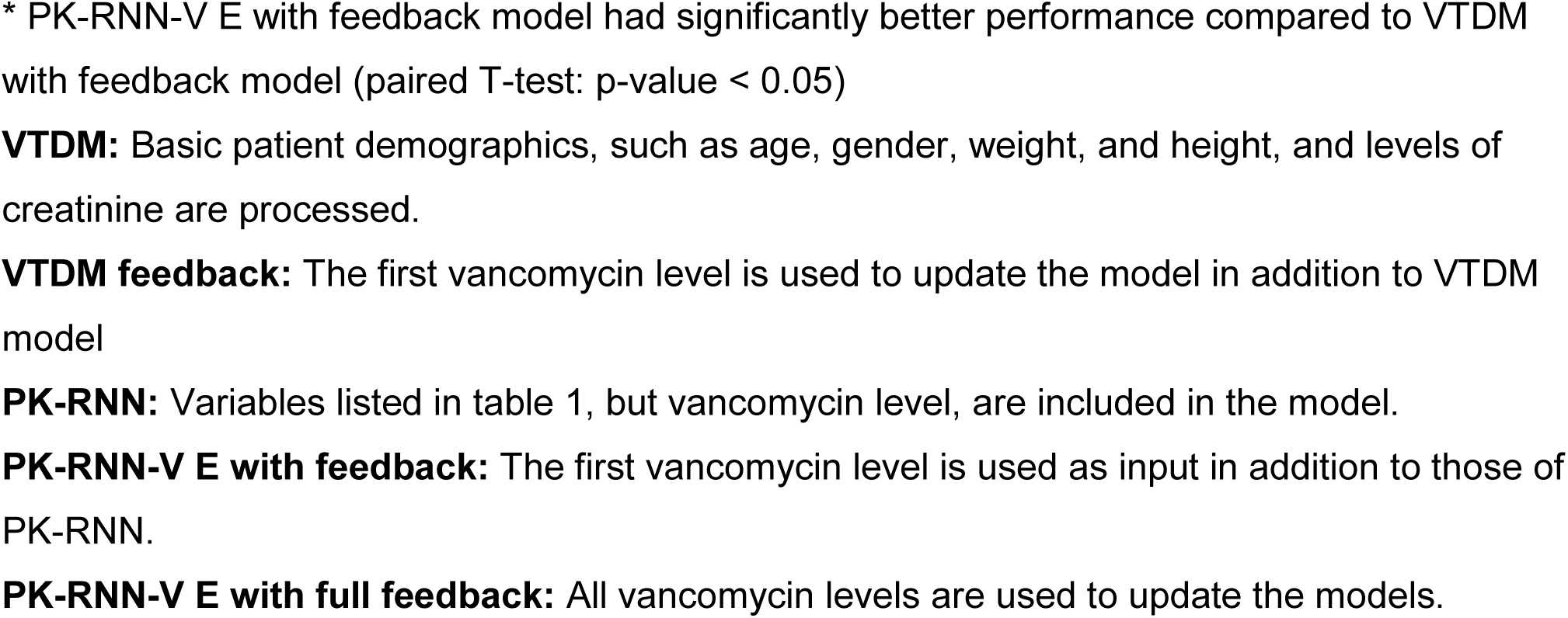
Model Performance Comparing Different Types of PK-RNN and Bayesian Models.

|  | RMSE | MAE | MAPE |
| --- | --- | --- | --- |
| **Bayesian Models** |  |  |  |
| VTDM | 8.58 | 6.54 | 41.81 |
| VTDM with Feedback | 6.29 | 4.26 | 29.15 |
| **PK-RNN-V models** |  |  |  |
| PK-RNN-V | 5.86 | 4.09 | 37.57 |
| PK-RNN-V E with Feedback | 5.39* | 3.64* | 25.41* |
| PK-RNN-V E with Full Feedback | 5.37 | 3.62 | 25.05 |

RMSE: Root mean square error, MAE: Mean absolute error, MAPE: Mean absolute percentage error, VTDM: Vancomycin Therapeutic Drug Model, RNN: Recurrent Neural Network, PK: Pharmacokinetics, 2477 encounters were included in the test dataset. The results for train and validation datasets are summarized in Supplementary Table 1.
\* PK-RNN-V E with feedback model had significantly better performance compared to VTDM with feedback model (paired T-test: $p$ -value < 0.05)
**VTDM:** Basic patient demographics, such as age, gender, weight, and height, and levels of creatinine are processed.
**VTDM feedback:** The first vancomycin level is used to update the model in addition to VTDM model
**PK-RNN:** Variables listed in table 1, but vancomycin level, are included in the model.
**PK-RNN-V E with feedback:** The first vancomycin level is used as input in addition to those of PK-RNN.
**PK-RNN-V E with full feedback:** All vancomycin levels are used to update the models.

The performance of the models in relation to the timing of vancomycin level measurements during the encounters also was evaluated (Table S2). PK-RNN-V performed better at predicting the first measurement of vancomycin levels, compared to VTDM (RMSE = 5.22 vs. 7.09, MAE = 3.87 vs. 4.85, MAPE = 32.95 vs. 65.18, respectively). Further, in regard to the second measurement of vancomycin levels, the PK-RNN-V E with feedback model outperformed the VTDM with feedback model with RMSE = 4.89 vs. 5.70 (*p*-value = 0.0022), MAE = 3.34 vs. 3.88 (*p*-value = 0.0022), and MAPE = 24.53 vs. 35.97 (*p*-value = 0.034). Finally, similar results were observed in the third measurement of vancomycin levels, except for MAPE, which did not reach statistical significance (RMSE = 5.94 vs. 6.94 [*p*-value = 0.00067], MAE = 4.00 vs. 4.72 [*p*-value = 0.00067], and MAPE = 28.59 vs. 44.64 [*p*-value = 0.12]). The lack of statistical significance is likely due to the number of patients who had three or more vancomycin measurements, which were limited; 435 patients vs. 222 patients were included in the second measurement groups and the three or more group, respectively.

Overall, PK-RNN-V models exhibited better performance across the subgroups, as shown in Table 4. PK-RNN-V models maintained their accuracy, even in patients with BMI > 35 (5.46 vs. 6.90, *p*-value = 0.0017, PK-RNN-V E with feedback and VTDM with feedback, respectively), age > 65 (4.53 vs. 5.47, p-value = 0.00063, PK-RNN-V E with feedback and VTDM with feedback, respectively), and patients on norepinephrine (5.47 vs. 5.84, *p*-value = 0.63, PK-RNN-V E with feedback and VTDM with feedback, respectively). All subgroups, except Hispanic patients and those on norepinephrine, had statistically significant results.

**Table 4.** Model Performance (RMSE) of Each Model in Different Patient Subgroups.

|  | Hispanic<br>N:389 | White<br>N:880 | AA<br>N:560 | Age<br>>65<br>N:925 | BMI<br>>30<br>N:982 | BMI<br>>35<br>N:551 | DM<br>N:671 | HTN<br>N:997 | CKD<br>N:297 | Norepi<sup>i</sup><br>N:123 |
| --- | --- | --- | --- | --- | --- | --- | --- | --- | --- | --- |
| **VTDM** | 8.21 | 7.65 | 9.66 | 7.72 | 8.92 | 9.64 | 7.75 | 8.04 | 8.3 | 11.93 |
| **VTDM with Feedback** | **6.04** | **5.25** | **7.62** | **5.47** | **6.3** | **6.9** | **5.47** | **5.21** | **5.44** | **5.84** |
| **PK-RNN-V** | 7.01 | 5.54 | 5.98 | 5.22 | 5.46 | 5.69 | 5.41 | 5.32 | 5.14 | 7.23 |
| **PK-RNN-V E with feedback** | **6.03** | **4.68\*** | **5.79\*** | **4.53\*** | **4.99\*** | **5.46\*** | **4.87\*** | **4.45\*** | **4.72\*** | **5.47** |
| **PK-RNN-V E with Full Feedback** | 5.92 | 4.66 | 5.88 | 4.48 | 4.95 | 5.41 | 4.81 | 4.42 | 4.7 | 5.44 |
| **PK-RNN-V E without Kidney Function** | 6.47 | 5.00 | 6.91 | 4.98 | 5.4 | 5.99 | 5.14 | 4.98 | 5.46 | 5.03 |

AA: African American, BMI: Body Mass Index, DM: Diabetes Mellitus, HTN: Hypertension, CKD: Chronic Kidney Diseases
<sup>i</sup> Patients who were on norepinephrine.
\* PK-RNN-V E with feedback model had significantly better performance compared to VTDM with feedback model (paired T-test: p-value < 0.05)

Figure 2 presents a sample of patients in our dataset who had better accuracy in each model (PK-RNN-V E and VTDM). Figure 2A presents the patient in whom the PK-RNN-V E with feedback model predicted the vancomycin levels better, whereas Figure 2B presents the opposite case. Each line in the upper figure is the predicted vancomycin level from each model and the patient’s demographics. The lower figure shows the changes in vancomycin dosage and creatinine levels over time. As shown in the figure, VTDM models have more steep PK curves as compared to PK-RNN-V E models. This is likely the difference between the one-compartment (PK-RNN-V E) and two-compartment (VTDM) models. In Figure 2A, PK-RNN-V E model predicted the first four vancomycin levels well. Despite actual vancomycin levels increased in the fourth and fifth measurements, the model accurately adjusted their curves to gain close predictions. The sixth label value was higher than the predicted values, whereas VTDM model already revealed bigger errors after the third prediction without adjustment of the curve. In Figure 2B, overall, both models predicted higher levels than actual values. Interestingly, both models slowly adjusted their curves, and the fourth and fifth curves are close to the actual values. Although our models need to be improved and validated before clinical use, those small errors in those patients are promising. Although we do not have gold standard AUC levels in our dataset, the RNN-PK-V E with a feedback model provided the AUC of vancomycin level based on the curves in Figure 2: mean AUC = 367.8, 372.8, and 364.5 in training, validation, and test datasets, respectively.

**Figure 2:**
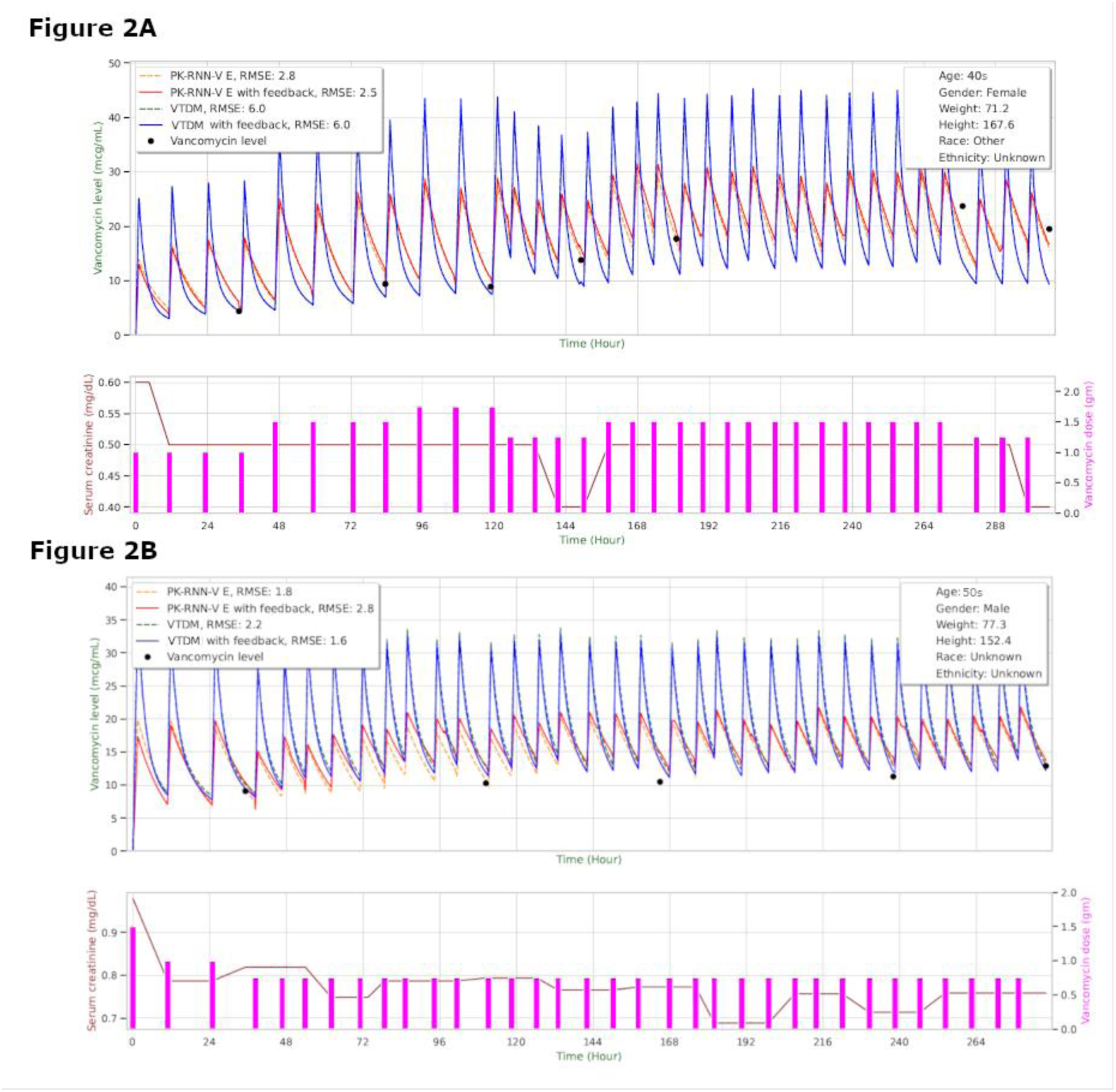
Predicted Vancomycin Serum Concentration Curves and Other Basic Characteristics in Sample Patients.

Each figure shows predicted vancomycin concentration curves, with actual vancomycin levels (black dots) in the upper figure, and the serum creatinine (solid purple line) and vancomycin administration and dose (purple bar) in the lower figure. The legend provides basic patient demographics (upper right corner) and RMSE of each model (upper left corner). The feedback models adjust the prediction based on the first vancomycin. **Figure 2A**. A sample patient in whom the PK-RNN-V E with feedback model predicted vancomycin levels better than did the VTDM model (Bayesian model). **Figure 2B**. A sample patient in whom the VTDM with feedback model (Bayesian model) predicted the vancomycin level better than did the PK-RNN-V E with feedback model.

## Discussion

The PK-RNN-V E model is a deep learning model used to forecast vancomycin serum concentrations in real time based on predicted personalized volume distribution (*v*) and elimination (*k*) of vancomycin from EHR data, including not only basic demographics and serum creatinine levels but also other patient-specific features, such as vital signs, laboratory results, and concomitant medications. The model demonstrated superior prediction of vancomycin levels, including trough and random levels, compared to the Bayesian model generated based on the publicly available models (VTDM model). Our model tolerated missing values in real-world data and offered accurate prediction across heterogeneous patient populations. The PK-RNN-V E model performed better predictions than did the VTDM model in the first vancomycin levels based on the data prior to the level (the first measurement of VTDM vs. PK-RNN-V E models, as seen in Table S2). This could provide an empirical vancomycin dosing recommendation as per the PK-RNN-V E model. Based on the predicted PK parameters, our model also conveniently generated the AUC/MIC (Figure 2) of vancomycin levels, which is currently part of the standard of care for vancomycin dosing.

Compared to Bayesian models, deep learning models, such as ours, are more expressive PK models, enabling the incorporation of dynamic, multimodal, and longitudinal data into the predictions. The model can disentangle the complex relationship among the large number of heterogeneous clinical inputs at irregular time steps. At each input time step in which some clinical measures or events take place, the RNN can update its internal state and predict PK parameters. Although our current model limited the time steps with vancomycin dosing, vancomycin levels, or the end of the day, a different time step schedule could be considered. This could be changed to a shorter time frame, for example, at the time of any lab work or any clinical events, such as vital sign measurement, with a minimal additional computational burden. Compared to the limited set of a patient’s characteristics that existing Bayesian models take to adjust the PK parameters, our PK-RNN-V E model can take into consideration broader and more complex patient-specific characteristics, as recorded in the real-world EHRs.

In some sense, both Bayesian models and PK-RNN models are models with sequential hidden variables and a passive PK model that generates outputs. The Bayesian models are limited in the capacity of their hidden variables, typically only those that are sufficient to specify the distribution of PK models. The PK-RNN models, however, can have a larger capacity (32-dimensional vector in our case), which not only encode the information needed to specify the PK models, but also with some “memory” that can carry through from timepoint to timepoint. In addition, PK-RNN models use arbitrary parameterized neural networks to specify the functional dependency from input variables to the hidden variables, and the hidden variables to the outputs, while Bayesian models are limited in certain families of the function as specified by the probability rules. This limitation of functional form of Bayesian models makes it difficult to incorporate complex real-world variables. However, the limit (“strong prior” on the functional form) of Bayesian models may make it more data-efficient when the training sample is small and the dependencies are relatively simple. For EHR data, we are in the large data regime with complex dependencies among variables, and thus PK-RNN models are more suited. In fact, Bayesian models can be seen as a special case of the PK-RNN models, if RNNs with a more complex functional form beyond simple one-layer gated linear layers in GRUs are used.

Our PK-RNN-V model can be expanded in several ways. Currently, our PK-RNN-V E model takes only limited structured data as proof of concept. Our previous work, however, finds that the deep learning models can be further advanced by adding more features or even unstructured data with minimal data processing.[27] Vancomycin does not have significant drug-drug interactions with other medications. Some medications, however, such as angiotensin (ACE) inhibitors, could affect patient creatinine levels, which is a key predictor of vancomycin eliminations in Bayesian models.[28] Deep learning models may be able to learn those interactions based on the data and predict the levels, taking these effects into consideration. We believe that this is particularly useful when the PK of medications is affected significantly by multiple drug-drug interactions. In current practice, although one-to-one drug-drug interaction can be evaluated based on the available PK data, the drug-drug interactions can be complicated when multiple concomitant medications possess interactions with each other.

By design, the PK-RNN-V E model can leverage the specific characteristics of the data from the local patient population and even the characteristics of the individual patient. We believe that the PK-RNN-V E model can provide more accurate and personalized levels. In fact, our model remained accurate in multiple patient subgroups. Further, our model maintained its accuracy even without creatinine levels or estimated GFRs in the dataset (masked models are found in the Supplementary Materials), which are aspects of the critical features to estimate vancomycin elimination in the Bayesian models. This highlights the power of deep learning models to predict vancomycin elimination with other features integrated into the model as “surrogate markers.” We also conducted multiple subgroups analyses. Overall, the PK-RNN-V E model exhibited better performance than did the Bayesian model, except for some subgroups, such as patients on norepinephrine (S Table 3). The limited number of patients on norepinephrine in our cohort hampered the training of the models and insufficiently powered the statistical analyses in the test dataset. The results warrant further studies in these specific populations.

For application of our models in clinical practice, there are several aspects that should be addressed. Recent studies have shown that maintaining the AUC/MIC of vancomycin in a certain range (between 400 and 600) predicts a better clinical response and could avoid nephrotoxicity from vancomycin as compared to trough-based targeted therapy.^3^ Because the actual AUC was not measured in our study, we did not compare the AUC of PK-RNN-V E and VTDM models and provided only the predicted AUC of PK-RNN-V E with feedback models. Future studies are warranted to address the accuracy of the AUC of PK-RNN-V E models. In addition, the recommended empirical dosage of vancomycin at initiation of the therapy was not calculated in our study. The dosage can be provided, however, using feedback from the first predicted vancomycin levels. Of note, our model provided a more accurate first vancomycin level than did the Bayesian model (Table S2), which provides evidence that the PK-RNN-V model could provide more individualized dosing recommendations even without using the first measurement of the vancomycin level. This potentially provides an advantage of being able to target vancomycin concentration/AUC faster in critically ill patients. In addition, our model could provide personalized vancomycin dosing at each administration based on the real-time data from EHRs, as clinical decision support systems in EHRs.

There are several limitations in our study. First, our study used retrospective data from a single healthcare system in Houston, Texas. Potential biases due to the study design could not be avoided. Although our healthcare system contains 14 inpatient hospitals, further studies are warranted to confirm our findings and the generalizability of the results. Second, we did not have access to commercially available Bayesian models for the comparison. VTDM models were created based on publicly available models and fine-tuned as the individualized model, as similar to commercially available models. There is a possibility, however, that those commercially available models may have better performance compared to VTDM models. Further, our model used a one-compartment model for vancomycin level prediction. Vancomycin PK can be characterized by a two- or multi-compartment model.[13] Although it is surprising that the one-compartment PK-RNN-V E model outperformed the two-compartment Bayesian model, PK-RNN-V E with multi-compartment models should be evaluated. Third, our study excluded patients who were receiving renal replacement therapy, such as hemodialysis or ECMO. Those patients, especially with those undergoing renal replacement therapy, have a different PK, requiring further model development and evaluation. Finally, as our PK-RNN-V E model integrates multiple features/variables from EHRs, compared to Bayesian models, the transferability and implementability of our model should be established for future applications in clinical use.

## Conclusion

Our deep learning-based model (PK-RNN-V E with feedback) provided better performance in predicting vancomycin levels against actual vancomycin levels as compared to the traditional Bayesian-based prediction model for vancomycin (VTDM with feedback) in a large retrospective real-world data set. These findings remained in subgroup analyses, which support the concepts and advantages of deep learning models. The PK-RNN-V E model can integrate real-time patient-specific data from an EHR, which allows real-time TDM and likely leads to precision dosing of vancomycin based on the TDM. Prospective and external validations of models are warranted in future studies.

## Data Availability

All data produced in the present study are available upon reasonable request to the authors.

## Abbreviations

AI: Artificial intelligence
ACE: Angiotensin
ASHP: American Society of Health-System Pharmacists
AUC: Area Under the Curve
BMI: Body Mass Index
CKD: Chronic Kidney Disease
DHHS: U.S. Department of Health and Human Services
DM: Diabetes Mellitus
ECMO: Extracorporeal Membrane Oxygenation
EHR: Electronic Health Record
HTN: Hypertension
ICU: Intensive Care Unit
IDSA: Infectious Diseases Society of America
IQR: Interquartile Range
IRB: Internal Review Boards
MAE: Mean Absolute Error
MAPE: Mean Absolute Percentage Error
MHHS: Memorial Hermann Health System
MIC: Minimal Inhibitory Concentration
MRSA: Methicillin resistant *Staphylococcus aureus*
SIDP: Society for Infectious Diseases Pharmacists
TDM: Therapeutic Drug Monitoring
PK: Pharmacokinetic
RMSE: Root Mean Square Error
RNN: Recurrent Neural Network
VAN: Vancomycin
VTDM: Vancomycin Therapeutic Drug Monitoring

## Acknowledgments

LR is supported by the UTHealth Innovation for Cancer Prevention Research Training Program Pre-Doctoral Fellowship (CPRIT Grant No. RP160015 and CPRIT Grant No. RP210042).

## Supplementary Materials

### 1. Detailed description of PK-RNN-V model

The 40 continuous variables in the input of PK-RNN-V model are following:

**S Table 1:** Variables Included in PK-RNN-V Model.

| Type of Features | Features |
| --- | --- |
| Time | Time (measured in days from the start of the encounter), Time elapsed since last event |
| Demographic data | Age at the time of visit, Gender, Ethnicity/Race, Height, Weight |
| Vital signs | Heart rate, Body temperature, Systolic blood pressure, Diastolic blood pressure, Respiratory rate, SpO2, O2 flow |
| Vancomycin dose | Vancomycin dose |
| Laboratory results | White blood cell count, Hematocrit, Hemoglobin, Red blood cell counts, Platelet counts, Neutrophils #, Lymphocytes (%), Lymphocytes counts, Monocytes (%), Monocytes counts, Eosinophils (%), Eosinophils counts, Basophils (%), Basophils counts, Nucleated red blood cell (%), Sodium level, Potassium level, Creatinine level, Blood urea nitrogen level, Estimated glomerular filtration rate, Bicarbonate level, Chloride level, Glucose level, Total calcium level, Phosphorus level, Magnesium level, Albumin level, Total bilirubin level, Total protein level |
| Additional features included in feedback models | Vancomycin concentration |

All of the lab measurements are normalized by subtracting the mean and dividing by the standard deviation. The features are updated when the values are updated.

The equation that we use to calculate the vancomycin concentration (*Vc*) is:

Vc = *M/Vd*, where *M* is the total mass of the vancomycin, and *Vd* is the volume of the distribution.

*M* is calculated as follows: *M*_*t*_ = *A* ∗ *M*_*t*−1_ + *B*, where *A* = *e*^−*k*∗*td*^ and 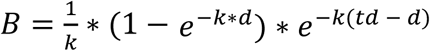. Here, *td* is the time difference between the current and previous events (in hours), and d is the duration of the infusion (in hours). We assume a constant vancomycin infusion rate of 1 gm/hr; thus, the value of *d* also is equal to the dose of vancomycin in grams. The variables *k* and *Vd* are generated from the recurrent neural network (RNN) model’s 32 dimensional hidden states by first applying a linear transform to reduce the dimensions to two, followed by an exponential transformation to make them non-negative.

The loss function of this model consists of four terms: the mean square error term and three regularizing terms, as described in the main text. The weight of the regularizing terms is 1e3. This weight is found through a grid search in the log scale, using the validation dataset.

### 2. Detailed description of the VTDM model

The traditional Bayesian VTDM model uses two compartments and has free parameters, *η*_1,_ *η*_2_, *η*_3_, that follow a Gaussian distribution: *η*_1_ ∼ *N*(0, 0.120), *η*_2_ ∼ *N*(0, 0.149), *η*_3_ ∼ *N*(0, 0.416), where the variances are estimated from another study. The vancomycin concentrations in the two compartments (blood and peripheral) are denoted by A and B, and the ordinary differential equations that describe the dynamics of A and B are

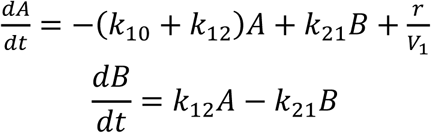

, where *k*_10_ indicates the elimination rate of serum vancomycin, *k*_12_ and *k*_21_ represent the exchange rate between two compartments, and *r* represents the infusion rate. The *η*s are related to the elimination and exchange rates *k* as per the following equations:

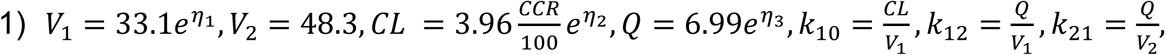

where the CCR is the clearance of creatinine calculated from the Cockcroft-Gault equation. Solving these ordinary differential equations, we get the serum vancomycin concentration as

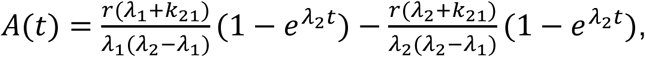

where 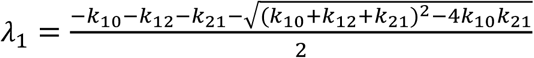 and 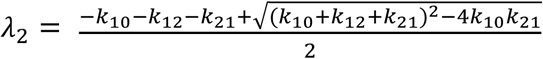. We first set all *η*s to be 0 and measure the prediction errors. We find that the mean difference between the first prediction and the measurement error is 4.85. Then we adjusted the *η*s based on the first measurement and used a gradient ascent to maximize the log-posterior 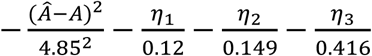 for each encounter separately, where *Â* is the first estimated concentration from the VTDM model, and *A* is the first measurement of concentration.

**S Table 2.** Variables Included in VTDM model.

STable 2 Variables Included in VTDM model
| Type of Features | Features |
| --- | --- |
| Time | Time (measured in days from the start of the encounter), Time elapsed since the last event |
| Demographic data | Age at the time of visit, Gender, Weight |
| Vancomycin dose | Vancomycin dose |
| Laboratory results | Creatinine level |
| Additional features included in feedback models | Vancomycin concentration |

### 3. Formulas of Cockcroft CrCl equation, RMSE, MAE, and MAPE

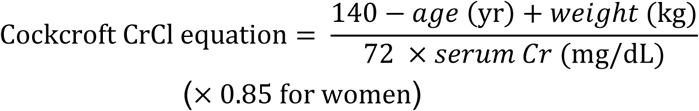

Metrics for “overall measurements”

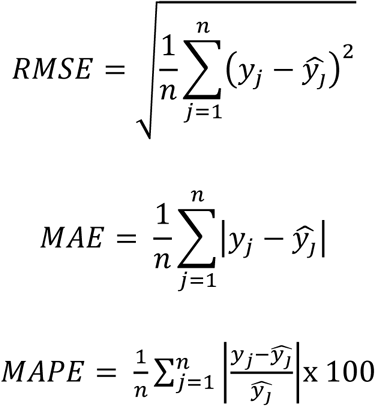

Metrics for “per encounter”

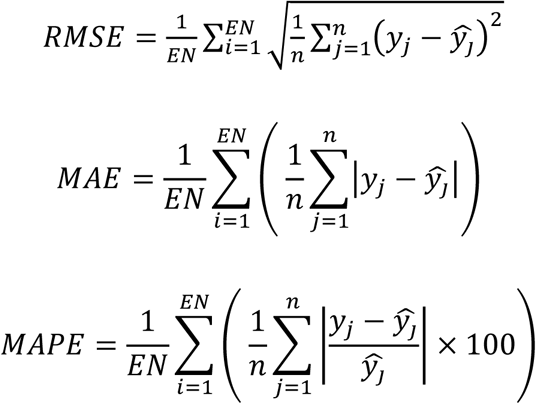

Metrics for “per patient”

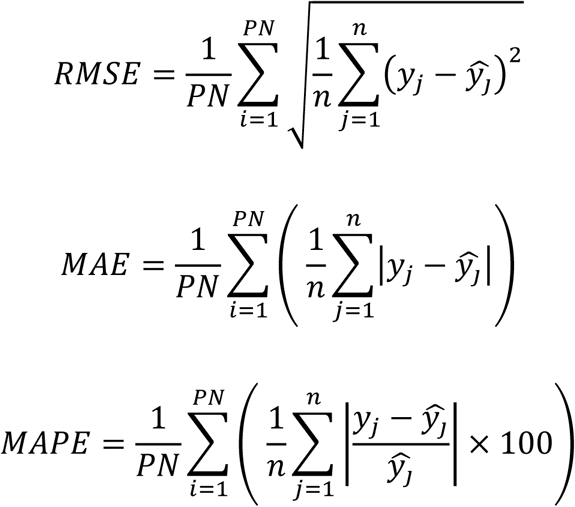

n: number of measurements, EN: number of encounters, PN: number of patients, *ŷ*^*j*^: predicted vancomycin level from the model, *y*_*j*_: actual vancomycin level

CrCl: Creatinine Clearance, RMSE: Root Mean Square Error, MAE: Mean Absolute Error, MAPE: Mean Absolute Percentage Error

**S Figure 1.**
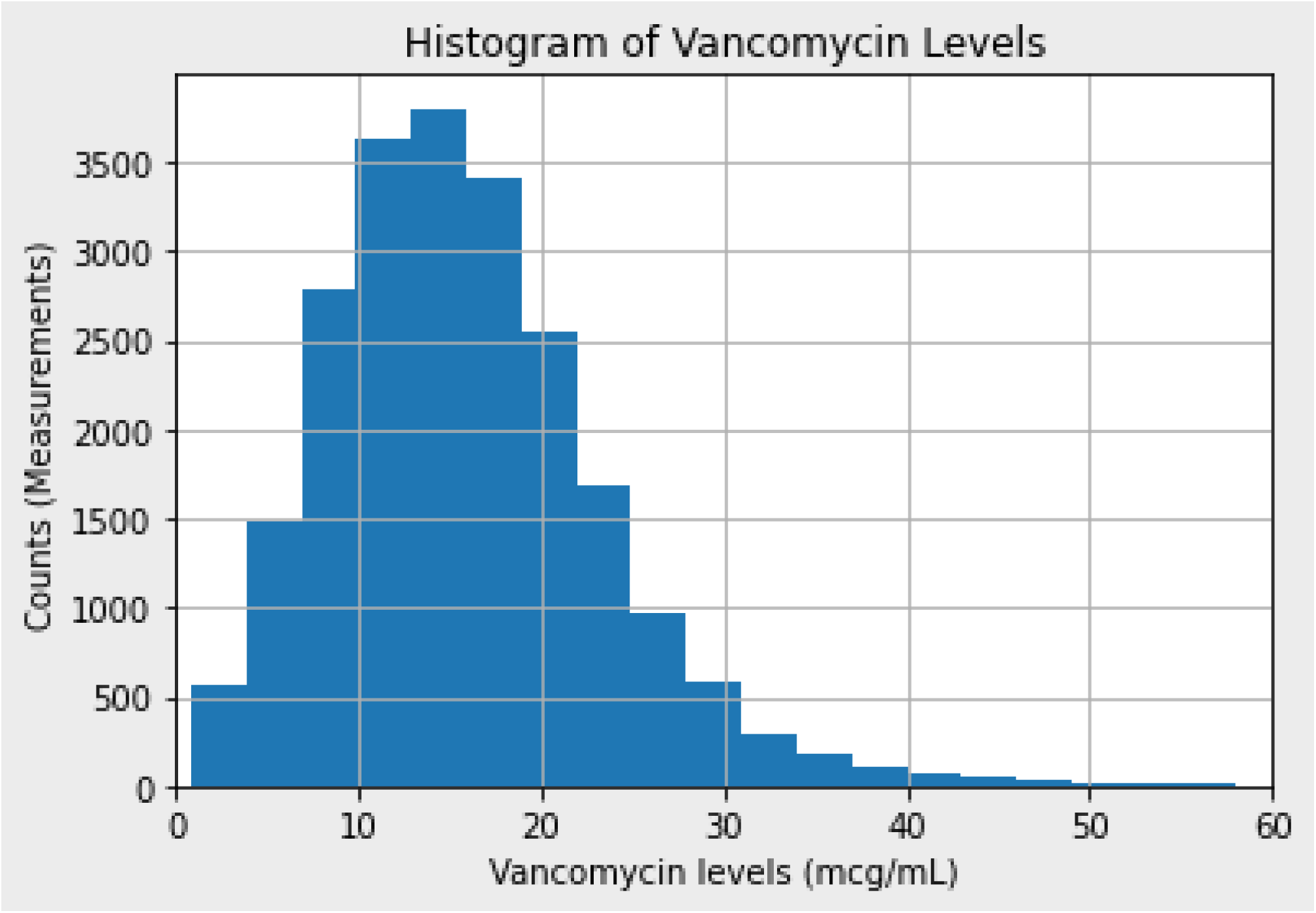
Histogram of Vancomycin Levels.

**S Table 3.** Accuracy based on Overall measurements, Encounter, and Patient in Train, Validation, and Test Datasets.

**5. S Table 3. Accuracy based on Overall measurements, Encounter, and Patient in Train, Validation, and Test Datasets**
|  |  | Train Data<br>Measurement: 11,240<br>Encounter: 6050<br>Patient: 3813 |  |  | Validation Data<br>Measurement: 2306<br>Encounter: 1250<br>Patient: 821 |  |  | Test Data<br>Measurement: 2438<br>Encounter: 1300<br>Patient: 806 |  |  |
| --- | --- | --- | --- | --- | --- | --- | --- | --- | --- | --- |
| Model | Type of Measurement | RMSE | MAE | MAPE | RMS E | MAE | MAPE | RMSE | MAE | MAPE |
| VTDM | Overall measurements | 8.09 | 6.16 | 38.73 | 7.24 | 5.5 | 35.24 | 8.58 | 6.54 | 41.81 |
|  | Per Encounter | 6.03 | 5.84 | 38.66 | 5.44 | 5.24 | 34.46 | 6.18 | 5.94 | 39.86 |
|  | Per Patient | 6.49 | 6.23 | 37.93 | 5.61 | 5.4 | 32.86 | 6.93 | 6.56 | 45.07 |
| VTDM with Feedback | Overall measurements | 6.07 | 4.3 | 28.08 | 5.27 | 3.82 | 24.81 | 6.29 | 4.26 | 29.55 |
|  | Per Encounter | 4.15 | 3.94 | 27.21 | 3.77 | 3.56 | 24.16 | 4.24 | 3.98 | 29.18 |
|  | Per Patient | 4.85 | 4.55 | 28.27 | 4.14 | 3.90 | 23.79 | 5.00 | 4.63 | 33.96 |
| PK-RNN-V | Overall measurements | 5.29 | 3.81 | 32.53 | 5.42 | 3.95 | 34.91 | 5.86 | 4.09 | 37.57 |
|  | Per Encounter | 4.00 | 3.74 | 32.46 | 4.18 | 3.93 | 35.68 | 4.18 | 3.92 | 38.83 |
|  | Per Patient | 4.21 | 3.80 | 31.87 | 4.08 | 3.76 | 35.97 | 4.49 | 4.01 | 34.62 |
| PK-RNN-V E with feedback | Overall measurements | 4.92 | 3.46 | 23.97 | 4.92 | 3.51 | 23.95 | **5.39\*** | **3.64\*** | **25.41\*** |
|  | Per Encounter | 3.48 | 3.25 | 23.63 | 3.66 | 3.42 | 23.48 | **3.61\*** | **3.37\*** | **24.32\*** |
|  | Per Patient | 3.81 | 3.51 | 23.25 | 3.66 | 3.43 | 22.19 | **4.20\*** | **3.80\*** | **24.03\*** |
| PK-RNN-V E with Full Feedback | Overall measurements | 4.86 | 3.42 | 23.81 | 4.87 | 3.47 | 23.52 | 5.37 | 3.62 | 25.05 |
|  | Per Encounter | 3.46 | 3.23 | 23.55 | 3.63 | 3.39 | 23.21 | 3.59 | 3.35 | 24.12 |
|  | Per Patient | 3.74 | 3.43 | 23.01 | 3.56 | 3.32 | 21.18 | 4.14 | 3.75 | 23.42 |
| PK-RNN-V E without Creatinine/GFR | Overall measurements | 5.19 | 3.72 | 26.16 | 5.43 | 3.98 | 27.34 | 5.91 | 4.01 | 27.14 |
|  | Per Encounter | 3.72 | 3.49 | 25.56 | 4.1 | 3.86 | 26.31 | 3.97 | 3.71 | 25.68 |
|  | Per Patient | 4.16 | 3.84 | 26.25 | 4.15 | 3.89 | 26.28 | 4.72 | 4.30 | 28.42 |

RMSE: Root mean square error, MAE: Mean absolute error, MAPE: Mean absolute percentage error, VTDM: Vancomycin Therapeutic Drug Model, RNN: Recurrent Neural Network, PK: Pharmacokinetics,
\* PK-RNN-V E with feedback model had significantly better performance compared to VTDM with feedback model (paired T-test: p-value < 0.05)
**VTDM:** Basic patient demographics, such as age, gender, weight, and height, and levels of creatinine are processed.
**VTDM feedback:** The first vancomycin level is used to update the model in addition to VTDM model
**PK-RNN:** Variables listed in table 1, but vancomycin level, are included in the model.
**PK-RNN-V E feedback:** The first vancomycin level is used in addition to PK-RNN.
**PK-RNN-V E full feedback:** All vancomycin levels are used to update the models.
**PK-RNN-V E without Creatinine/GFR:** Creatinine levels and GFR were excluded from predicting features.

**S Table 4.** All Model Accuracy Results Depending on the Timing of Vancomycin Level Measurements.

6. S Table 4. All Model Accuracy Results Depending on the Timing of Vancomycin Level Measurements
|  |  | First Measurement<br>Measurement: 1300<br>Encounter: 1300<br>Patient: 806 |  |  | Second Measurement<br>Measurement: 622<br>Encounter: 622<br>Patient: 453 |  |  | Third or More Measurements<br>Measurement: 516<br>Encounter: 277<br>Patient: 222 |  |  |
| --- | --- | --- | --- | --- | --- | --- | --- | --- | --- | --- |
| Model | Type of Measurement | RMSE | MAE | MAPE | RMSE | MAE | MAPE | RMSE | MAE | MAPE |
| **VTDM** | **Overall measurements** | 7.09 | 4.85 | 65.18 | 7.77 | 5.9 | 90.88 | 9.47 | 7.31 | 135.59 |
|  | **Per Encounter** | 4.85 | 4.85 | 65.18 | 5.9 | 5.9 | 90.88 | 6.83 | 6.59 | 117.61 |
|  | **Per Patient** | 4.73 | 4.49 | 57.1 | 5.74 | 5.57 | 76.24 | 6.93 | 6.56 | 100.37 |
| **VTDM with Feedback** | **Overall measurements** | n/a | n/a | n/a | 5.7 | 3.88 | 35.97 | 6.94 | 4.72 | 44.64 |
|  | **Per Encounter** | n/a | n/a | n/a | 3.88 | 3.88 | 35.97 | 4.81 | 4.56 | 43.85 |
|  | **Per Patient** | n/a | n/a | n/a | 3.85 | 3.68 | 33.72 | 5.00 | 4.63 | 42.28 |
| **PK-RNN-V** | **Overall measurements** | 5.22 | 3.87 | 32.95 | 5.62 | 3.95 | 39.63 | 6.14 | 4.26 | 35.08 |
|  | **Per Encounter** | 3.87 | 3.87 | 32.95 | 3.95 | 3.95 | 39.63 | 4.25 | 3.96 | 38.28 |
|  | **Per Patient** | 4.04 | 3.80 | 33.22 | 4.10 | 3.93 | 38.90 | 4.32 | 3.92 | 35.92 |
| **PK-RNN-V E with Feedback** | **Overall measurements** | n/a | n/a | n/a | **4.89\*** | **3.34\*** | **24.53\*** | **5.94\*** | **4.00\*** | **28.59** |
|  | **Per Encounter** | n/a | n/a | n/a | **3.34\*** | **3.34\*** | **24.53\*** | **4.08\*** | **3.80\*** | **29.04** |
|  | **Per Patient** | n/a | n/a | n/a | **3.34\*** | **3.21\*** | **24.94\*** | **4.20\*** | **3.80\*** | **29.90** |
| **PK-RNN-V E with Full Feedback** | **Overall measurements** | n/a | n/a | n/a | 4.89 | 3.34 | 24.53 | 5.9 | 3.96 | 28.94 |
|  | **Per Encounter** | n/a | n/a | n/a | 3.34 | 3.34 | 24.53 | 4.01 | 3.74 | 28.98 |
|  | **Per Patient** | n/a | n/a | n/a | 3.34 | 3.21 | 24.94 | 4.14 | 3.75 | 29.95 |

RMSE: Root Mean Square Error, MAE: Mean Absolute Error, MAPE: Mean Absolute Percentage Error
n/a: The results were excluded as feedback model uses the first measurement to completely fit the model to the actual values before the accuracy measurements.
\* PK-RNN-V E with feedback model had significantly better performance compared to VTDM with feedback model (paired T-test: p-value < 0.05)

**7. S Table 5.** Model Accuracy in Different Subgroups. S Table 5-A. Model Accuracy by “Overall measurements” in Different Subgroups

a. Root Mean Square Error (RMSE)
|  | **Hispanic**<br>**N:389** | **White**<br>**N:880** | **AA**<br>**N:560** | **Age**<br>**>65**<br>**N:925** | **BMI**<br>**>30**<br>**N:982** | **BMI**<br>**>35**<br>**N:551** | **DM**<br>**N:671** | **HTN**<br>**N:997** | **CKD**<br>**N:297** | **Norepi<sup>i</sup>**<br>**N:123** |
| --- | --- | --- | --- | --- | --- | --- | --- | --- | --- | --- |
| **VTDM** | 8.21 | 7.65 | 9.66 | 7.72 | 8.92 | 9.64 | 7.75 | 8.04 | 8.3 | 11.93 |
| **VTDM with Feedback** | **6.04** | **5.25** | **7.62** | **5.47** | **6.3** | **6.9** | **5.47** | **5.21** | **5.44** | **5.84** |
| **PK-RNN-V** | 7.01 | 5.54 | 5.98 | 5.22 | 5.46 | 5.69 | 5.41 | 5.32 | 5.14 | 7.23 |
| **PK-RNN-V E with feedback** | **6.03** | **4.68\*** | **5.79\*** | **4.53\*** | **4.99\*** | **5.46\*** | **4.87\*** | **4.45\*** | **4.72\*** | **5.47** |
| **PK-RNN-V E with Full Feedback** | 5.92 | 4.66 | 5.88 | 4.48 | 4.95 | 5.41 | 4.81 | 4.42 | 4.7 | 5.44 |
| **PK-RNN-V E without Kidney Function** | 6.47 | 5.00 | 6.91 | 4.98 | 5.4 | 5.99 | 5.14 | 4.98 | 5.46 | 5.03 |

**b. Mean Absolute Error (MAE)**
|  | Hispanic<br>N:389 | White<br>N:880 | AA<br>N:560 | Age<br>>65<br>N:925 | BMI<br>>30<br>N:982 | BMI<br>>35<br>N:551 | DM<br>N:671 | HTN<br>N:997 | CKD<br>N:297 | Norepi<sup>i</sup><br>N:123 |
| --- | --- | --- | --- | --- | --- | --- | --- | --- | --- | --- |
| **VTDM** | 6.12 | 5.78 | 7.29 | 6 | 7.15 | 7.91 | 6.06 | 6.38 | 6.53 | 9.63 |
| **VTDM with Feedback** | **4.14** | **3.77** | **4.76** | **3.9** | **4.33** | **4.74** | **3.97** | **3.92** | **4.14** | **4.08** |
| **PK-RNN-V** | 4.84 | 4.01 | 3.78 | 3.86 | 4.03 | 4.25 | 3.94 | 3.88 | 4.00 | 5.21 |
| **PK-RNN-V E with feedback** | **3.84** | **3.35\*** | **3.61\*** | **3.36\*** | **3.64\*** | **3.89\*** | **3.47\*** | **3.36\*** | **3.52\*** | **4.23** |
| **PK-RNN-V E with Full Feedback** | 3.78 | 3.34 | 3.62 | 3.32 | 3.62 | 3.88 | 3.44 | 3.31 | 3.49 | 4.24 |
| **PK-RNN-V E without Kidney Function** | 4.22 | 3.7 | 4.32 | 3.68 | 3.97 | 4.34 | 3.69 | 3.74 | 4.12 | 4.00 |

**c. Mean Absolute Percentage Error (MAPE)**
|  | Hispani<br>c<br>N:389 | White<br>N:880 | AA<br>N:560 | Age<br>>65<br>N:925 | BMI<br>>30<br>N:982 | BMI<br>>35<br>N:551 | DM<br>N:671 | HTN<br>N:997 | CKD<br>N:297 | Norepi<sup>i</sup><br>N:123 |
| --- | --- | --- | --- | --- | --- | --- | --- | --- | --- | --- |
| **VTDM** | 47.11 | 35.84 | 41.98 | 34.9 | 42.67 | 46.63 | 36.44 | 38.1 | 33.57 | 51.08 |
| **VTDM with Feedback** | **37.18** | **23.92** | **30.08** | **22.77** | **28.33** | **31.23** | **26.04** | **24.76** | **22.07** | **22.94** |
| **PK-RNN-V** | 49.39 | 30.11 | 29.84 | 32.34 | 43.31 | 37.78 | 39.92 | 37.45 | 31.08 | 37.60 |
| **PK-RNN-V E with Feedback** | **25.27** | **23.61** | **24.59\*** | **21.50** | **25.21\*** | **27.93\*** | **22.45\*** | **20.98\*** | **20.17** | **24.61** |
| **PK-RNN-V E with Full Feedback** | 24.46 | 23.36 | 24.49 | 21.11 | 24.87 | 27.61 | 22.12 | 20.67 | 19.97 | 24.69 |
| **PK-RNN-V E without Kidney Function** | 28.96 | 25.76 | 28.52 | 23.54 | 26.6 | 29.1 | 24.52 | 24.46 | 27.48 | 23.48 |

AA: African American, BMI: Body Mass Index, DM: Diabetes Mellitus, HTN: Hypertension, CKD: Chronic Kidney Diseases
<sup>i</sup> Patients who were on norepinephrine.
\* PK-RNN-V E with feedback model had significantly better performance compared to VTDM with feedback model (paired T-test: p-value < 0.05) In this table, only test dataset results are summarized.

**S Table 5B.**
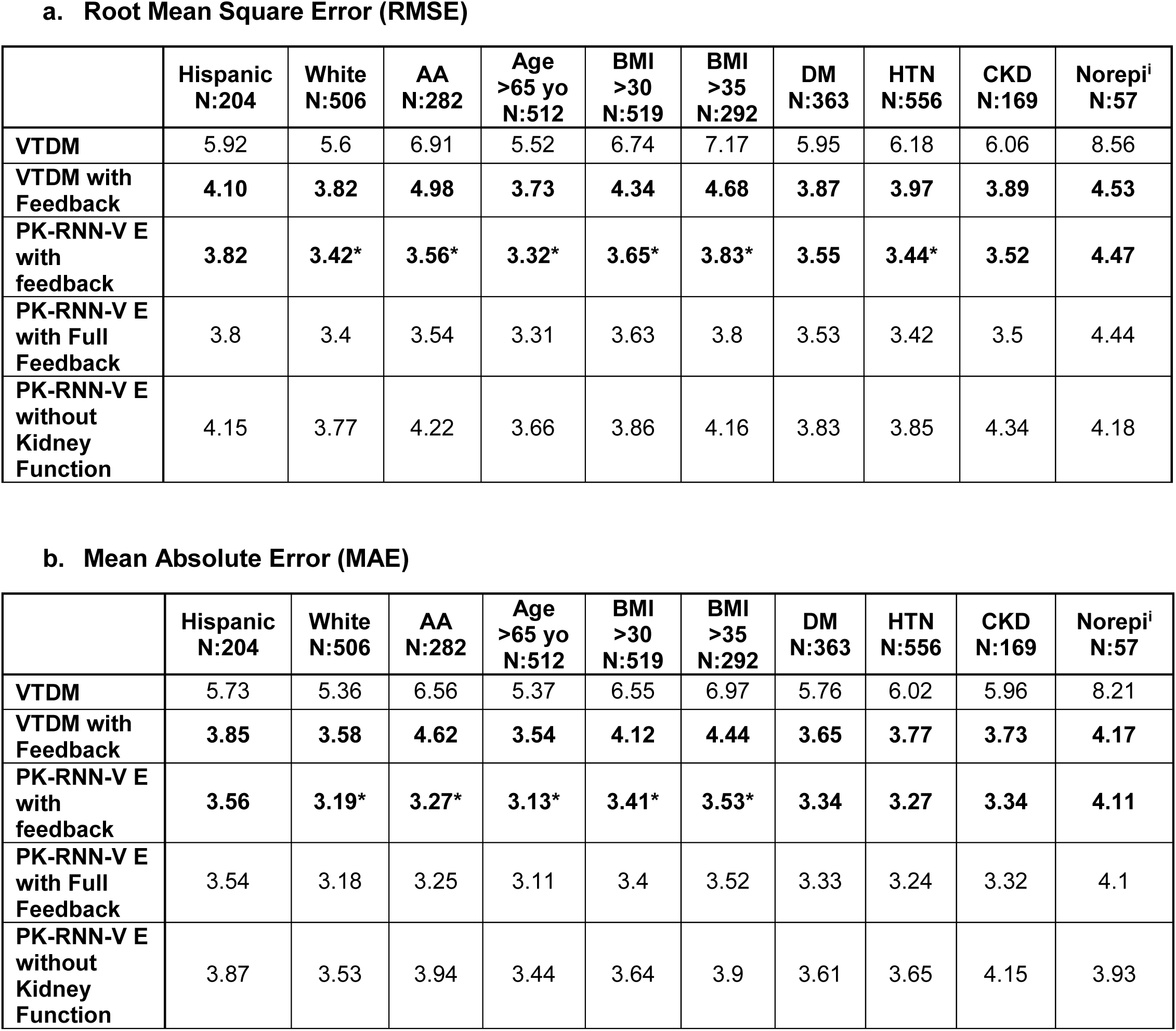
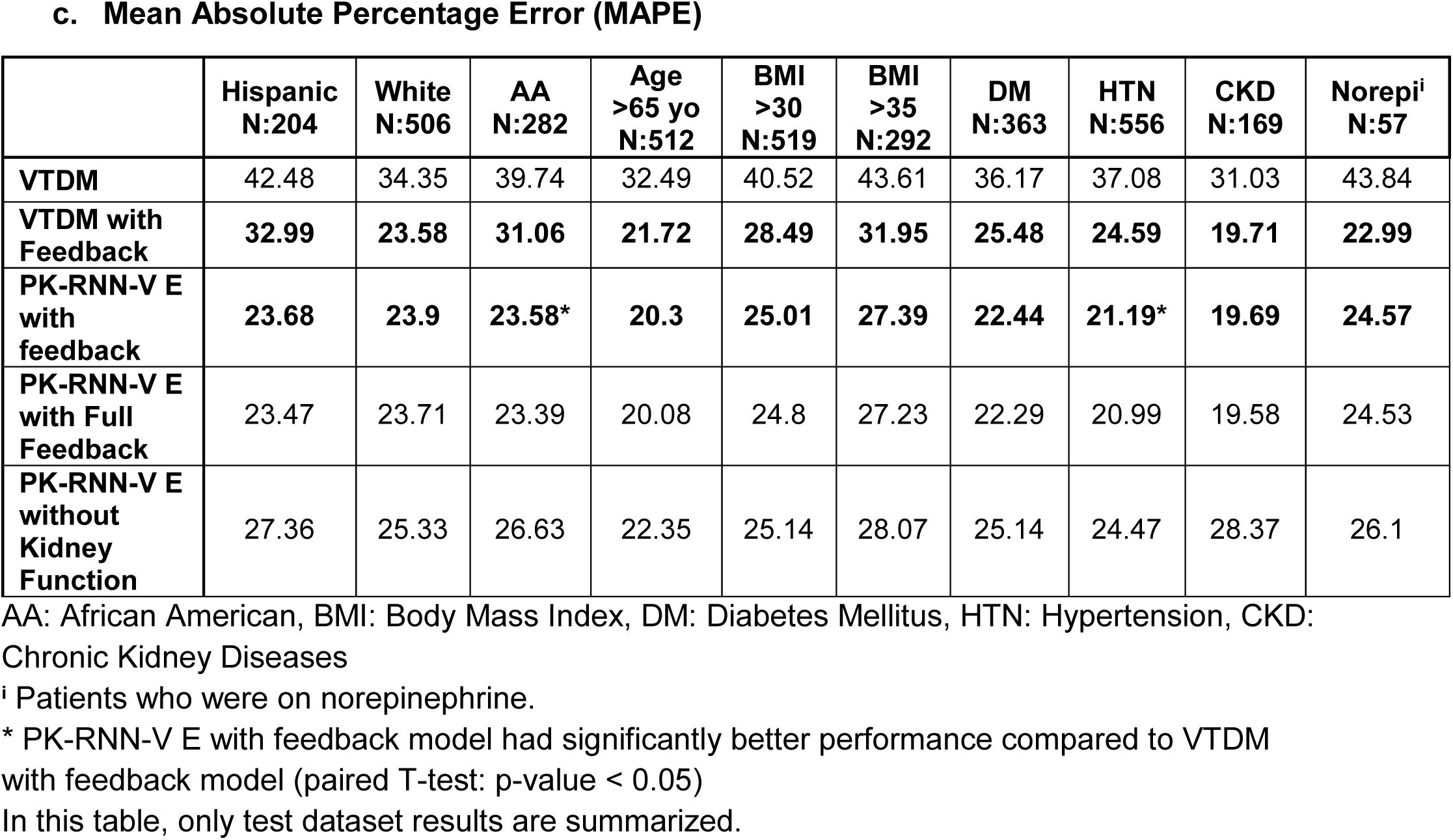
Model Accuracy by “per encounter” in Different Subgroups.

**S Table 5 B. Model Accuracy by “per encounter” in Different Subgroups** **a. Root Mean Square Error (RMSE)**
|  | Hispanic<br>N:204 | White<br>N:506 | AA<br>N:282 | Age<br>>65 yo<br>N:512 | BMI<br>>30<br>N:519 | BMI<br>>35<br>N:292 | DM<br>N:363 | HTN<br>N:556 | CKD<br>N:169 | Norepi<sup>i</sup><br>N:57 |
| --- | --- | --- | --- | --- | --- | --- | --- | --- | --- | --- |
| **VTDM** | 5.92 | 5.6 | 6.91 | 5.52 | 6.74 | 7.17 | 5.95 | 6.18 | 6.06 | 8.56 |
| **VTDM with Feedback** | **4.10** | **3.82** | **4.98** | **3.73** | **4.34** | **4.68** | **3.87** | **3.97** | **3.89** | **4.53** |
| **PK-RNN-V E with feedback** | **3.82** | **3.42\*** | **3.56\*** | **3.32\*** | **3.65\*** | **3.83\*** | **3.55** | **3.44\*** | **3.52** | **4.47** |
| **PK-RNN-V E with Full Feedback** | 3.8 | 3.4 | 3.54 | 3.31 | 3.63 | 3.8 | 3.53 | 3.42 | 3.5 | 4.44 |
| **PK-RNN-V E without Kidney Function** | 4.15 | 3.77 | 4.22 | 3.66 | 3.86 | 4.16 | 3.83 | 3.85 | 4.34 | 4.18 |

**b. Mean Absolute Error (MAE)**
|  | Hispanic<br>N:204 | White<br>N:506 | AA<br>N:282 | Age<br>>65 yo<br>N:512 | BMI<br>>30<br>N:519 | BMI<br>>35<br>N:292 | DM<br>N:363 | HTN<br>N:556 | CKD<br>N:169 | Norepi<sup>i</sup><br>N:57 |
| --- | --- | --- | --- | --- | --- | --- | --- | --- | --- | --- |
| **VTDM** | 5.73 | 5.36 | 6.56 | 5.37 | 6.55 | 6.97 | 5.76 | 6.02 | 5.96 | 8.21 |
| **VTDM with Feedback** | **3.85** | **3.58** | **4.62** | **3.54** | **4.12** | **4.44** | **3.65** | **3.77** | **3.73** | **4.17** |
| **PK-RNN-V E with feedback** | **3.56** | **3.19\*** | **3.27\*** | **3.13\*** | **3.41\*** | **3.53\*** | **3.34** | **3.27** | **3.34** | **4.11** |
| **PK-RNN-V E with Full Feedback** | 3.54 | 3.18 | 3.25 | 3.11 | 3.4 | 3.52 | 3.33 | 3.24 | 3.32 | 4.1 |
| **PK-RNN-V E without Kidney Function** | 3.87 | 3.53 | 3.94 | 3.44 | 3.64 | 3.9 | 3.61 | 3.65 | 4.15 | 3.93 |

**c. Mean Absolute Percentage Error (MAPE)**
|  | Hispanic<br>N:204 | White<br>N:506 | AA<br>N:282 | Age<br>>65 yo<br>N:512 | BMI<br>>30<br>N:519 | BMI<br>>35<br>N:292 | DM<br>N:363 | HTN<br>N:556 | CKD<br>N:169 | Norepi<sup>i</sup><br>N:57 |
| --- | --- | --- | --- | --- | --- | --- | --- | --- | --- | --- |
| **VTDM** | 42.48 | 34.35 | 39.74 | 32.49 | 40.52 | 43.61 | 36.17 | 37.08 | 31.03 | 43.84 |
| **VTDM with Feedback** | **32.99** | **23.58** | **31.06** | **21.72** | **28.49** | **31.95** | **25.48** | **24.59** | **19.71** | **22.99** |
| **PK-RNN-V E with feedback** | **23.68** | **23.9** | **23.58\*** | **20.3** | **25.01** | **27.39** | **22.44** | **21.19\*** | **19.69** | **24.57** |
| **PK-RNN-V E with Full Feedback** | 23.47 | 23.71 | 23.39 | 20.08 | 24.8 | 27.23 | 22.29 | 20.99 | 19.58 | 24.53 |
| **PK-RNN-V E without Kidney Function** | 27.36 | 25.33 | 26.63 | 22.35 | 25.14 | 28.07 | 25.14 | 24.47 | 28.37 | 26.1 |

AA: African American, BMI: Body Mass Index, DM: Diabetes Mellitus, HTN: Hypertension, CKD: Chronic Kidney Diseases
<sup>i</sup> Patients who were on norepinephrine.
\* PK-RNN-V E with feedback model had significantly better performance compared to VTDM with feedback model (paired T-test: p-value < 0.05)
In this table, only test dataset results are summarized.

**S Table 5C.** Model Accuracy by “per patient” in Different Subgroups.

**a. Root Mean Square Error (RMSE)**
|  | Hispanic<br>N:131 | White<br>N:316 | AA<br>N:154 | Age<br>>65 yo<br>N:350 | BMI<br>>30<br>N:362 | BMI<br>>35<br>N:203 | DM<br>N:194 | HTN<br>N:324 | CKD<br>N:98 | Norepi<sup>i</sup><br>N:52 |
| --- | --- | --- | --- | --- | --- | --- | --- | --- | --- | --- |
| **VTDM** | 5.78 | 6.22 | 8.07 | 6.3 | 7.5 | 8.4 | 6.14 | 6.69 | 6.5 | 8.9 |
| **VTDM with Feedback** | **4.50** | **4.30** | **5.88** | **4.69** | **4.91** | **5.6** | **4.43** | **4.58** | **4.46** | **5.21** |
| **PK-RNN-V E with feedback** | **4.54\*** | **3.89\*** | **4.4\*** | **3.94\*** | **3.96\*** | **4.48\*** | **3.58\*** | **3.85\*** | **4.21** | **5.95** |
| **PK-RNN-V E with Full Feedback** | 4.5 | 3.86 | 4.33 | 3.87 | 3.93 | 4.46 | 3.55 | 3.79 | 4.18 | 5.88 |
| **PK-RNN-V E without Kidney Function** | 5.18 | 4.25 | 5.24 | 4.46 | 4.51 | 5.04 | 3.98 | 4.26 | 4.92 | 4.96 |

**b. Mean Absolute Error (MAE)**
|  | Hispanic<br>N:131 | White<br>N:316 | AA<br>N:154 | Age<br>>65 yo | BMI<br>>30 | BMI<br>>35 | DM<br>N:194 | HTN<br>N:324 | CKD<br>N:98 | Norepi<sup>i</sup><br>N:52 |
| --- | --- | --- | --- | --- | --- | --- | --- | --- | --- | --- |

|  |  |  |  | N:350 | N:362 | N:203 |  |  |  |  |
| --- | --- | --- | --- | --- | --- | --- | --- | --- | --- | --- |
| **VTDM** | 5.45 | 5.86 | 7.44 | 6.02 | 7.19 | 8.11 | 5.76 | 6.31 | 6.14 | 8.43 |
| **VTDM with Feedback** | **4.17** | **3.9** | **5.19** | **4.35** | **4.52** | **5.16** | **4.01** | **4.16** | **4.00** | **4.70** |
| **PK-RNN-V E with feedback** | **4.07\*** | **3.54\*** | **3.80\*** | **3.64\*** | **3.56\*** | **3.99\*** | **3.11\*** | **3.46\*** | **3.77** | **5.38** |
| **PK-RNN-V E with Full Feedback** | 4.06 | 3.52 | 3.75 | 3.57 | 3.55 | 3.98 | 3.1 | 3.4 | 3.74 | 5.34 |
| **PK-RNN-V E without Kidney Function** | 4.74 | 3.9 | 4.67 | 4.14 | 4.13 | 4.62 | 3.57 | 3.88 | 4.48 | 4.55 |

c. Mean Absolute Percentage Error (MAPE)
|  | Hispanic<br>N:131 | White<br>N:316 | AA<br>N:154 | Age<br>>65 yo<br>N:350 | BMI<br>>30<br>N:362 | BMI<br>>35<br>N:203 | DM<br>N:194 | HTN<br>N:324 | CKD<br>N:98 | Norepi<sup>i</sup><br>N:52 |
| --- | --- | --- | --- | --- | --- | --- | --- | --- | --- | --- |
| **VTDM** | 61.63 | 37.12 | 48.74 | 34.65 | 43.26 | 47.1 | 41.01 | 43.33 | 31.44 | 41.85 |
| **VTDM with Feedback** | **54.56** | **24.46** | **37.64** | **24.66** | **28.00** | **30.17** | **32.35** | **30.9** | **21.19** | **23.16** |
| **PK-RNN-V E with feedback** | **27.2** | **21.99** | **25.23\*** | **21.36\*** | **23.27\*** | **25.78\*** | **20.52\*** | **21.49\*** | **22.59** | **27.1** |
| **PK-RNN-V E with Full Feedback** | 26.83 | 21.6 | 24.78 | 20.69 | 23.02 | 25.5 | 20.39 | 21.07 | 22.38 | 26.95 |
| **PK-RNN-V E without Kidney Function** | 35.03 | 26.11 | 31.66 | 26.62 | 26.94 | 29.71 | 25.78 | 26.57 | 33.66 | 23.77 |

AA: African American, BMI: Body Mass Index, DM: Diabetes Mellitus, HTN: Hypertension, CKD: Chronic Kidney Diseases
<sup>i</sup> Patients who were on norepinephrine.
\* PK-RNN-V E with feedback model had significantly better performance compared to VTDM with feedback model (paired T-test: p-value < 0.05)
In this table, only test dataset results are summarised.

**8. S Table 6.** Diagnostic Codes used for Baseline Characteristics.

| **Comorbidities** | **ICD 9 or 10 codes** |
| --- | --- |
| Cerebral vascular diseases | I60-I69, 430-438 |
| Coronary artery diseases | I25.10, I25.83, Z95.5, I25.810, 414.01, V45.82 |
| Heart failure | 398.91, 402.01, 402.11, 402.91, 404.01, 404.03, 404.11, 404.13, 404.91, 404.93, I09.9, I11.0, I13.0, I13.2, I25.5, I42.0, P29.0, I50.32, I50.23, I42.5-I42.9, 425.4-425.9, I43.x, I50.x, 428.x |
| Chronic pulmonary diseases | I27.8, I27.9, J68.4, J70.1, J70.3, 506.4, 416.8, 416.9, J44.0, J44.1, J84.9, 508.1, 508.8, J98.01, 518.1, 770.2, 748.61, 516.31, 500, 502, 503, 505, Z99.81, V46.2, J40.x-J47.x, J60.x-J70.x, 490.x-505.x, J84.x, J98.x |
| Hypertension | 401.1, 401.9, 642.0, I10, I13.0, I10.x, 401.x, 402.10, 402.90, 404.10, 404.90, 405.1, 405.9, 401.0, 642.1, 642.2, 642.7, 642.9, I13.2, I13.10, I13.11, I12.9, I13.0, I11.0, I13.0, 402.x-405.x, I11.x-I13.x, I15.x |
| Diabetes | 648.0, E10.0, E10.1, E10.9, E11.0, E11.1, E11.9, E12.0, E12.1, E12.9, E13.0, E13.1, E13.9, E14.0, E14.1, E14.9, 250.0-250.3, 775.1, E11.10, 250.4-250.9, E10.2-E10.8, E11.2-E11.8, E12.2-E12.8, E13.2-E13.8, E14.2-E14.8 |
| Chronic kidney diseases (CKD) | 403.11, 403.91, 404.12, 404.92, V42.0, V45.1, V56.0, V56.8, 403.01, 404.02, 404.03, 404.13, 404.93, I12.0, I13.1, N25.0, Z94.0, Z199.2, 588.0, I13.0, 585.x, 586.x, V56.x, N18.x, N19.x, Z49.0-Z49.2 |
| Hemodialysis | N18.6, Z99.2, 585.6, 39.95, 54.98, V45.1 |
| HIV/AIDS | B20.x-B22.x, B24.x, 042.x-044.x |
| Cancer | 203.8, 238.6, 273.3, V10.71, V10.72, V10.79, C90.0, C90.2, 203.0, 200.x-202.3x, 202.5-203.0, C81.x-C85.x, C88.x, C96.x, 200.x-202.x, 196.x-199.x, C77.x-C80.x, 140.x-172.x, 174.x, 175.x, 179.x-195.x, V10.x, C00.x-C26.x, C30.x-C34.x, C37.x-C41.x, C43.x, C45.x-C58.x, C60.x-C76.x, C97.x, 174.x-195.x |
| Solid Organ Transplantation | V42.9, Z94.0, V42.0, Z94.4, V42.7, Z94.2, V42.6, Z94.1, V42.1, V42.2, V42.83, Z94.83, V42.84, Z94.82 |

ECMO status was retrieved from nurse documentation such as ECMO settings.
HIV: Human immunodeficiency virus, AIDS: Acquired Immunodeficiency syndrome

**9. S Table 7.**
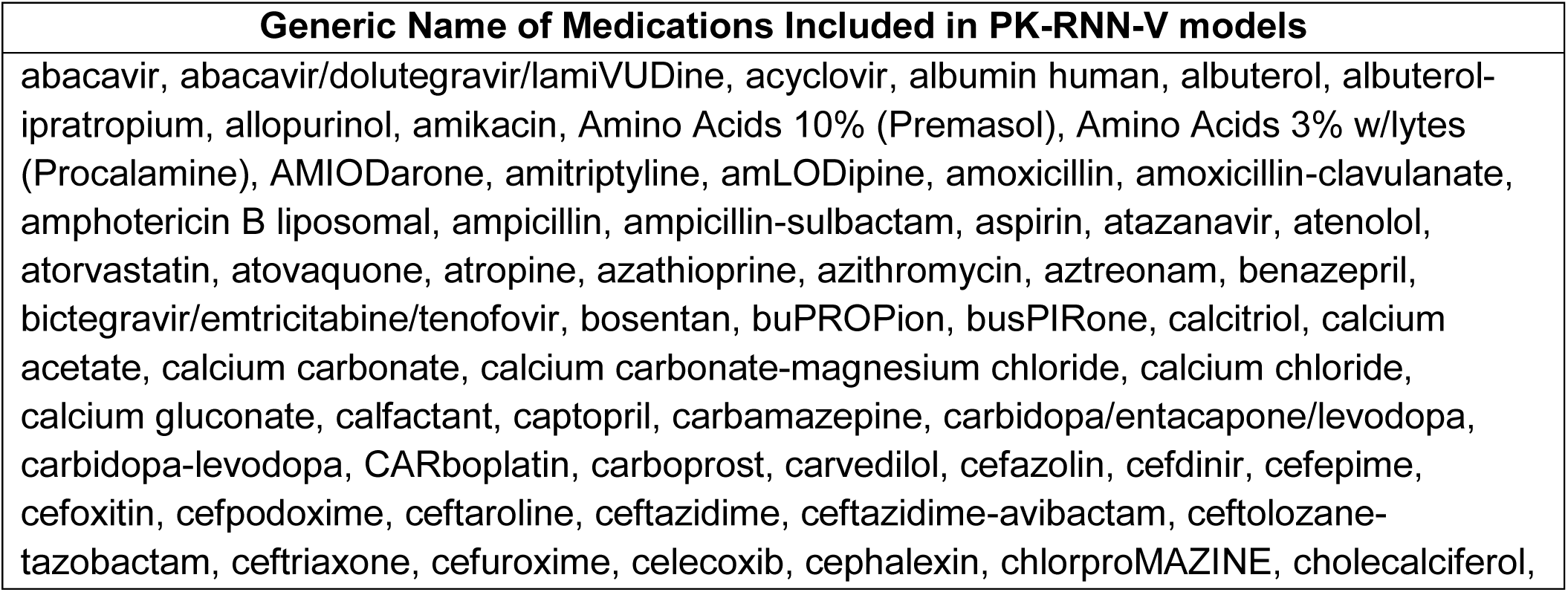
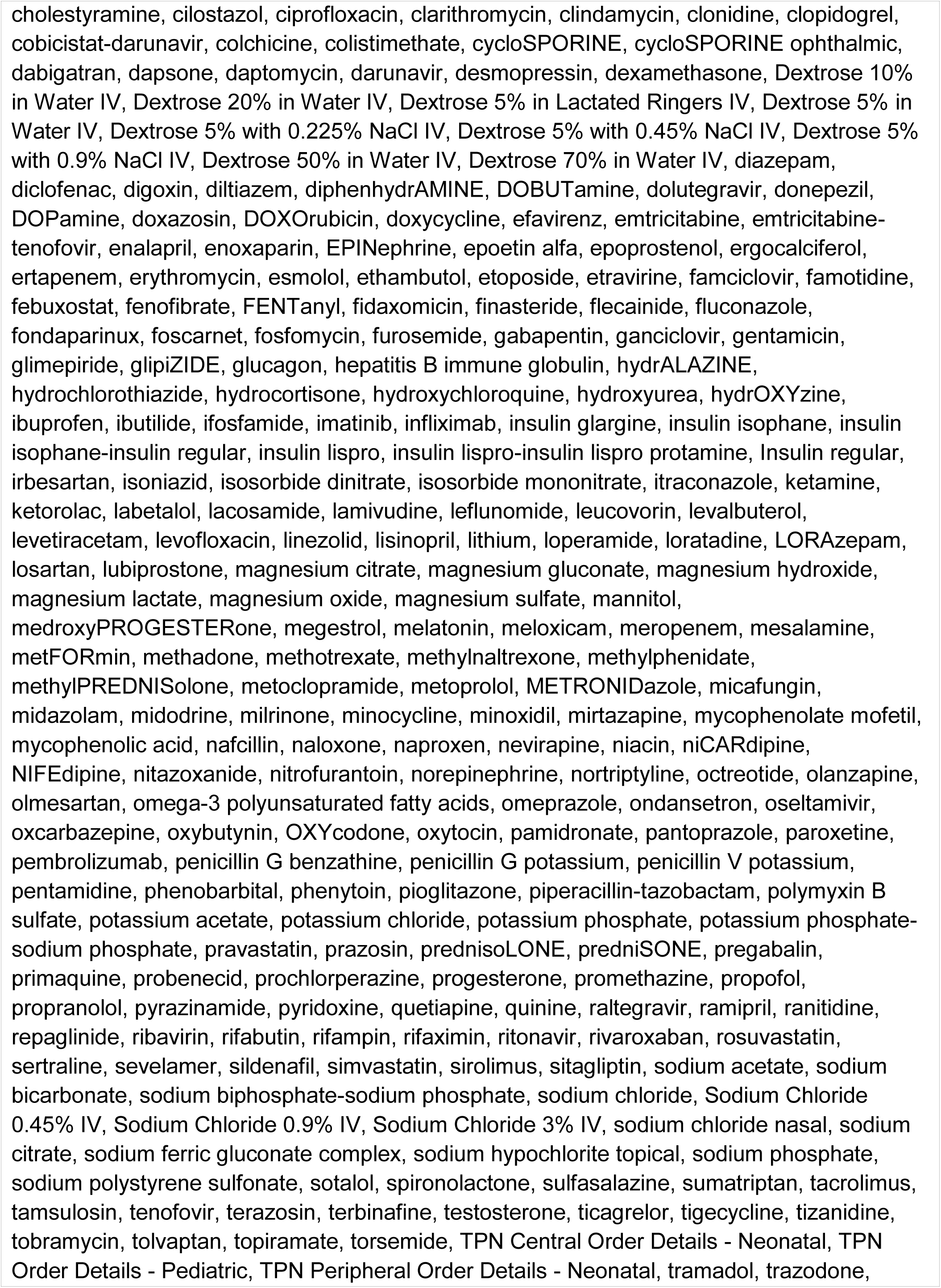
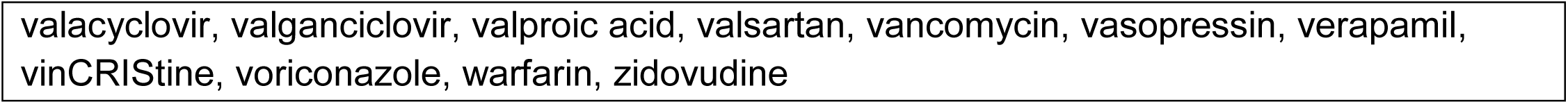
Generic Name of Medications Included in PK-RNN-V models.

## References

[1] M.J. Rybak, J.C. Rotschafer, K.A. Rodvold, Vancomycin: Over 50 Years Later and Still a Work in Progress, Pharmacotherapy: The Journal of Human Pharmacology and Drug Therapy. 33 (2013) 1253–1255. https://doi.org/10.1002/phar.1382.

[2] H.F. Chambers, F.R. DeLeo, Waves of Resistance: Staphylococcus aureus in the Antibiotic Era, Nat Rev Microbiol. 7 (2009) 629–641. https://doi.org/10.1038/nrmicro2200.

[3] M.J. Rybak, J. Le, T.P. Lodise, D.P. Levine, J.S. Bradley, C. Liu, B.A. Mueller, M.P. Pai, A. Wong-Beringer, J.C. Rotschafer, K.A. Rodvold, H.D. Maples, B.M. Lomaestro, Therapeutic monitoring of vancomycin for serious methicillin-resistant Staphylococcus aureus infections: A revised consensus guideline and review by the American Society of Health-System Pharmacists, the Infectious Diseases Society of America, the Pediatric Infectious Diseases Society, and the Society of Infectious Diseases Pharmacists, American Journal of Health-System Pharmacy. 77 (2020) 835–864. https://doi.org/10.1093/ajhp/zxaa036.

[4] M.L. Avent, V.L. Vaska, B.A. Rogers, A.C. Cheng, S.J. van Hal, N.E. Holmes, B.P. Howden, D.L. Paterson, Vancomycin therapeutics and monitoring: a contemporary approach, Internal Medicine Journal. 43 (2013) 110–119. https://doi.org/10.1111/imj.12036.

[5] S.W. Narayan, Y. Thoma, P.G. Drennan, H. Yejin Kim, J.-W. Alffenaar, S. Van Hal, A.E. Patanwala, Predictive Performance of Bayesian Vancomycin Monitoring in the Critically Ill, Critical Care Medicine. Publish Ahead of Print (2021). https://doi.org/10.1097/CCM.0000000000005062.

[6] D.S. Buelga, M. del Mar Fernandez de Gatta, E.V. Herrera, A. Dominguez-Gil, M.J. García, Population pharmacokinetic analysis of vancomycin in patients with hematological malignancies, Antimicrob Agents Chemother. 49 (2005) 4934–4941. https://doi.org/10.1128/AAC.49.12.4934-4941.2005.

[7] A. Broeker, M. Nardecchia, K.P. Klinker, H. Derendorf, R.O. Day, D.J. Marriott, J.E. Carland, S.L. Stocker, S.G. Wicha, Towards precision dosing of vancomycin: a systematic evaluation of pharmacometric models for Bayesian forecasting, Clinical Microbiology and Infection. 25 (2019) 1286.e1-1286.e7. https://doi.org/10.1016/j.cmi.2019.02.029.

[8] C.Y. Spong, D.W. Bianchi, Improving Public Health Requires Inclusion of Underrepresented Populations in Research, JAMA. 319 (2018) 337–338. https://doi.org/10.1001/jama.2017.19138.

[9] K. Maxfield, I. Zineh, Precision Dosing: A Clinical and Public Health Imperative, JAMA. 325 (2021) 1505–1506. https://doi.org/10.1001/jama.2021.1004.

[10] C. Deng, T. Liu, K. Wu, S. Wang, L. Li, H. Lu, T. Zhou, D. Cheng, X. Zhong, W. Lu, Predictive performance of reported population pharmacokinetic models of vancomycin in Chinese adult patients, Journal of Clinical Pharmacy and Therapeutics. 38 (2013) 480–489. https://doi.org/10.1111/jcpt.12092.

[11] Z. Lin, D. Chen, Y.-W. Zhu, Z. Jiang, K. Cui, S. Zhang, L. Chen, Population pharmacokinetic modeling and clinical application of vancomycin in Chinese patients hospitalized in intensive care units, Scientific Reports. 11 (2021) 2670. https://doi.org/10.1038/s41598-021-82312-2.

[12] Adoption of Electronic Health Record Systems among U.S. Non-Federal Acute Care Hospitals: 2008-2015, (n.d.). https://dashboard.healthit.gov/evaluations/data-briefs/non-federal-acute-care-hospital-ehr-adoption-2008-2015.php (accessed August 8, 2021).

[13] M.J. Rybak, The Pharmacokinetic and Pharmacodynamic Properties of Vancomycin, Clin Infect Dis. 42 (2006) S35–S39. https://doi.org/10.1086/491712.

[14] B. Shickel, P.J. Tighe, A. Bihorac, P. Rashidi, Deep EHR: A Survey of Recent Advances in Deep Learning Techniques for Electronic Health Record (EHR) Analysis, IEEE J Biomed Health Inform. 22 (2018) 1589–1604. https://doi.org/10.1109/JBHI.2017.2767063.

[15] O. for C. Rights (OCR), Methods for De-identification of PHI, HHS.Gov. (2012). https://www.hhs.gov/hipaa/for-professionals/privacy/special-topics/de-identification/index.html (accessed April 13, 2020).

[16] Y. Bengio, R. Ducharme, P. Vincent, C. Jauvin, A Neural Probabilistic Language Model, (n.d.) 19.

[17] T.B. Brown, B. Mann, N. Ryder, M. Subbiah, J. Kaplan, P. Dhariwal, A. Neelakantan, P. Shyam, G. Sastry, A. Askell, S. Agarwal, A. Herbert-Voss, G. Krueger, T. Henighan, R. Child, A. Ramesh, D.M. Ziegler, J. Wu, C. Winter, C. Hesse, M. Chen, E. Sigler, M. Litwin, S. Gray, B. Chess, J. Clark, C. Berner, S. McCandlish, A. Radford, I. Sutskever, D. Amodei, Language Models are Few-Shot Learners, ArXiv:2005.14165 [Cs]. (2020). http://arxiv.org/abs/2005.14165 (accessed June 26, 2021).

[18] A.N. Jagannatha, H. Yu, Bidirectional RNN for Medical Event Detection in Electronic Health Records, Proc Conf. 2016 (2016) 473–482. https://doi.org/10.18653/v1/n16-1056.

[19] E. Choi, A. Schuetz, W.F. Stewart, J. Sun, Using recurrent neural network models for early detection of heart failure onset, J Am Med Inform Assoc. 24 (2017) 361–370. https://doi.org/10.1093/jamia/ocw112.

[20] F. Ma, R. Chitta, J. Zhou, Q. You, T. Sun, J. Gao, Dipole: Diagnosis Prediction in Healthcare via Attention-based Bidirectional Recurrent Neural Networks, Proceedings of the 23rd ACM SIGKDD International Conference on Knowledge Discovery and Data Mining. (2017) 1903–1911. https://doi.org/10.1145/3097983.3098088.

[21] L. Rasmy, Y. Wu, N. Wang, X. Geng, W.J. Zheng, F. Wang, H. Wu, H. Xu, D. Zhi, A study of generalizability of recurrent neural network-based predictive models for heart failure onset risk using a large and heterogeneous EHR data set, Journal of Biomedical Informatics. 84 (2018) 11–16. https://doi.org/10.1016/j.jbi.2018.06.011.

[22] S.S. Jambhekar, P.J. Breen, Basic Pharmacokinetics, 1st edition, Pharmaceutical Press, London, 2009.

[23] E.M.P. Widmark, Studies in the concentration of indifferent narcotics in blood and tissues, Acta Medica Scandinavica. 52 (1919) 87–164. https://doi.org/10.1111/j.0954-6820.1919.tb08277.x.

[24] J.E. Murphy, D.E. Gillespie, C.V. Bateman, Predictability of vancomycin trough concentrations using seven approaches for estimating pharmacokinetic parameters, American Journal of Health-System Pharmacy. 63 (2006) 2365–2370. https://doi.org/10.2146/ajhp060047.

[25] H.-S. Lim, Y.P. Chong, Y.-H. Noh, J.-A. Jung, Y.S. Kim, Exploration of optimal dosing regimens of vancomycin in patients infected with methicillin-resistant Staphylococcus aureus by modeling and simulation, J Clin Pharm Ther. 39 (2014) 196–203. https://doi.org/10.1111/jcpt.12123.

[26] asancpt/shiny-vtdm, Asan CPT, 2021. https://github.com/asancpt/shiny-vtdm (accessed June 1, 2021).

[27] L. Rasmy, M. Nigo, B.S. Kannadath, Z. Xie, B. Mao, K. Patel, Y. Zhou, W. Zhang, A. Ross, H. Xu, D. Zhi, Recurrent neural network models (CovRNN) for predicting outcomes of patients with COVID-19 on admission to hospital: model development and validation using electronic health record data, Lancet Digit Health. (2022) S2589-7500(22)00049–8. https://doi.org/10.1016/S2589-7500(22)00049-8.

[28] E. Andreev, M. Koopman, L. Arisz, A rise in plasma creatinine that is not a sign of renal failure: which drugs can be responsible?, Journal of Internal Medicine. 246 (1999) 247–252. https://doi.org/10.1046/j.1365-2796.1999.00515.x.

